# Independent associations of sleep timing, duration and quality with adiposity and weight status in a national sample of adolescents: the UK Millennium Cohort Study

**DOI:** 10.1101/2021.02.04.21249454

**Authors:** Paul J Collings

## Abstract

**Background:** There is evidence that short sleep elevates obesity risk in youth, but sleep is a multidimensional construct and few studies have investigated parameters beyond duration.

**Objectives:** To investigate if sleep onset time, duration, latency, and night waking frequency are independently associated with adiposity and weight status in adolescents.

**Methods:** This was a cross-sectional observational study of 10,619 13-15y olds who provided self-reported sleep characteristics and underwent an anthropometric assessment to determine adiposity (body mass index (BMI) z-score and percent body fat (%BF)) and weight status. Adjusted linear and logistic regressions were used to investigate associations.

**Results:** Compared to a sleep onset time before 10pm, later sleep was associated with higher adiposity and higher likelihood of overweight and obesity in boys (after midnight, odds ratio (95% CI): 1.76 (1.19 to 2.60), *p*=0.004) and girls (between 11-11:59pm: 1.36 (1.17 to 1.65), *p*=0.002). Compared to sleeping for >9-10 hours, sleeping for ≤8 hours was associated with higher likelihood of overweight and obesity (boys: 1.80 (1.38 to 2.35), *p*<0.001); girls: 1.38 (1.06 to 1.79), *p*=0.016); there was evidence of a U-shaped association in girls for whom >10 hours of sleep was also associated with higher likelihood of overweight and obesity (1.31 (1.06 to 1.62); *p*=0.014). In girls, relative to a sleep latency of 16-30 minutes, sleep latencies ≥46 minutes were associated with higher adiposity (46-60 minutes, %BF: 1.47 (0.57 to 2.36), *p*=0.001) and higher likelihood of overweight and obesity (46-60 minutes: 1.39 (1.05 to 1.83); *p*=0.020). Often as opposed to never waking in the night was also associated with higher adiposity in girls (BMI z-score: 0.24 (0.08 to 0.41), *p*=0.004; %BF: 1.44 (0.44 to 2.44), *p*=0.005).

**Conclusions:** Sleep duration and timing, and sleep quality in girls, are independently associated with adiposity and weight status in adolescence and may be important targets for obesity prevention.

## Introduction

Developmental shifts in sleep physiology predispose adolescents to biologically driven evening alertness. Around the same time parental enforcement of bedtimes typically fade, academic pressures increase, social networks expand, and sedentary screen-based activities which are stimulating or produce light stimuli increasingly take place late into evenings; these factors reinforce evening alertness and perpetuate a cycle of late sleep onset [1]. When accompanied by imposed early mornings for school insufficient sleep and tiredness result; both are prevalent in adolescents worldwide [2, 3]. This has far-reaching ramifications for health and well-being, including repercussions for adiposity levels and obesity risk [4]. A plethora of studies have investigated short sleep relative to weight-for-height indices or weight status categories. Meta-analysed results indicate that short sleep elevates obesity risk in youth [5].

Sleep is a multidimensional construct of partly overlapping dimensions, and it is increasingly recognised that that parameters such as sleep timing and quality may be influential with regards to health [6]. A small number of studies have found that later bed and sleep onset times are associated with higher body mass index (BMI) z-score [7, 8], waist circumference and percent body fat [8], overweight and obesity [7, 9], and obesity severity in adolescents [10]. Sleep quality encompasses an array of continuity and architecture features [11]. A recent meta-analysis found that poor sleep quality, which was defined broadly as longer sleep latency, more sleep disturbances, recurrent night waking, or lower sleep efficiency, predicted 46% (95% confidence interval (CI): 1.24 to 1.72) higher odds of overweight and obesity in young subjects [12]. It has consistently been shown that associations between short sleep duration with higher adiposity are stronger in boys than girls [4, 5]. Whether or not this may partly be due to sex differences in other sleep characteristics remains unknown. The few studies conducted to date on sleep timing and quality have been conducted in small and select samples, and effect modification by sex has not been investigated. This is an important knowledge gap to fill as adolescent girls experience poorer sleep quality than boys [13]. Whereas duration may be more tightly related to adiposity levels in boys, sleep quality could be a stronger determinant of adiposity in girls. Information of this type could help to optimise obesity prevention policies and strategies, by highlighting that sleep components should be tailored according to sex-specific requirements.

In addition it is important to decipher the possible pathways by which sleep parameters may relate to adiposity. The best available evidence remains of low quality, but suggests that biological and behavioural factors likely underpin the associations of sleep characteristics with adiposity. Inadequate sleep may stimulate hormone-mediated preferences for nutritionally poor calorific foodstuffs, which could be compounded by poor sleep enabling more time to eat, shifting meal times to later in the day, creating more opportunities for screen-based sedentary time, and reducing daily physical activity due to fatigue [14, 15]. Providing additional evidence to support one or more of the hypothesised pathways could help to ameliorate the consequences of inadequate sleep by identifying modifiable intermediate behaviours that can be targeted for mitigation. A research focus on adolescence is important, it is key transitional stage of growth and maturation that is characterised by marked behavioural changes in sleep, diet, sedentary behaviour and physical activity. It is a critical period for the development of persistent obesity [16].

This study examined the associations of multiple sleep dimensions, including sleep timing, duration and quality (sleep latency and night waking), with BMI *z*-score, percent body fat, and overweight and obesity in a large and nationally representative sample of UK adolescents. Sleep dimensions were mutually adjusted for one another and effect modification by sex was examined. To shed light on pathways of action, associations were scrutinised when adjusted for potential mediating factors including diet, screen time, and habitual physical activity.

## Methods

### Study population

The Millennium Cohort Study (MCS) is a nationally representative birth cohort study of children born in the UK between 2000 and 2002. A total of 18,818 children were recruited to the cohort [17]. A sixth sweep of data collection was conducted in 2015-16, when children were followed-up in adolescence and for the first time provided a detailed description of their sleeping habits [18]. This is a complete case analysis included only the first child surveyed in any household (thereby excluding a small number of twin and triplet siblings (*n*=121)) who aged 13-15y provided full data for sleep, adiposity, and potentially important covariates.

### Sleep characteristics

Adolescents chose one from five options to indicate when they usually went to sleep on school nights and non-school nights (“Before 9pm” (8:30) / “9-9:59pm” (9:30) / “10-10:59pm” (10:30) / “11-11:59pm” (11:30) / “After midnight” (12:30)). Average sleep onset times were calculated using weekday to weekend weighting in the ratio 5:2 and were collapsed to four groups (Before 10pm (assigned the reference group) / 10-10:59pm / 11-11:59pm / After midnight). Sleep onset times were combined with usual wake-up times on school days (“Before 6am” (5:30) / “6-6:59am” (6:30) / “7-7:59am” (7:30) / “8-8:59am” (8:30) / “After 9am” (9:30)) and non-school days (“Before 8am” (7:30) / “8-8:59am” (8:30) / “9-9:59am” (9:30) / “10-10:59am” (10:30) / “11-11:59am” (11:30) / “After midday” (12:30)) to calculate average sleep durations, which were weighted weekdays to weekends in the ratio 5:2 and were collapsed to four groups (≤8 hours / >8-9 hours / >9-10 hours (reference) / >10 hours). Adolescents reported how many minutes in the last four weeks it had usually taken them to fall asleep (0-15 / 16-30 (reference) / 31-45 / 46-60 / >60) and how often in the last four weeks they had woken in the night and had trouble getting back to sleep (Never (reference) / A little / Sometimes / Often / Most of the time / All of the time); the final two categories were combined to ‘Habitually’.

### Adiposity and weight status

Trained researchers followed standard protocols to measure weight (Tanita BF-522W) and height (Leicester height measure). Body mass index (BMI) was calculated and converted to *z*-scores and weight status categories; underweight and normal weight categories were combined [19]. Body fat percentage was predicted from height, weight and demographic factors using a validated prediction model [20]. Body fat percentage was also estimated by bioelectrical impedance analysis (Tanita BF-522W), but as this method can be influenced by numerous environmental and physical factors, and because some participants (*n*=135) opted out of the assessment, bioimpedance data were used only in a sensitivity analysis [21].

### Covariables

Potential covariables were selected based on evidence linking them with sleep and observations in the MCS cohort that they predict adolescent weight status [22, 23]. Age was calculated as the elapsed time between the child’s date of birth and the date of the sixth sweep assessment; which was also used to infer season of measurement. Adolescents self-reported their ethnic group [24] and parents reported their incomes which were equivalised to indicate net disposable household income per week [25]. Birth weight and the child’s breast feeding history were also parent reported. Adolescents reported the number of days in the last week they had accumulated at least an hour of moderate to vigorous physical activity (None / 1-2 days / 3-4 days / 5-6 days / Every day). They also reported the number of hours on a normal week day that they (a) watched programmes or films on TV, DVD, computer or mobile devices, (b) spent on social networking or messaging sites or Apps on the internet, and (c) spent playing electronic games on a computer or games system (None / <0.5h / 0.5-0.99h / 1-1.99h / 2-2.99h / 3-4.99h / 5-6.99h / ≥7h). A diet index (ranging from 0-10 with higher scores indicating healthier diet) was developed from five items covering the frequency that adolescents consumed (a) breakfast, (b) at least two portions of fruit per day, (c) at least two portions of vegetables per day, (d) sugary drinks, and (e) fast food; for the first three items three response options were available (Never (0) / Sometimes (1) / Every day (2)) whereas for sugary drinks and fast food seven original options were collapsed to three to derive the index (≥2 days per week (0) / 1-2 days per week (1) / Never or less than once a month (2)).

### Statistics

Linear regression was used to investigate associations of sleep characteristics with adiposity and logistic regression quantified associations with weight status. Initial models adjusted associations for age, ethnicity, net disposal household income, birth weight, breast feeding history, and season of measurement. Model 1 further adjusted for what could be perceived as potential confounding or mediating factors, including physical activity, TV viewing, and the healthy diet index. Model 3 adjusted sleep onset and duration for sleep latency and night waking frequency, and vice versa; sleep latency and night waking frequency were also mutually adjusted for each other. In sensitivity analyses, adjustment for TV viewing was replaced by adjustment for social media use or electronic gaming, and body fat percentage estimated by bioimpedance was modelled as an outcome. Analyses were performed with Stata/SE 16.1 software (StataCorp, College Station, TX), sex interactions were investigated using multiplicative terms, and survey weights were applied throughout [26]. *A priori p*<0.05 was deemed statistically significant but the results are interpreted with emphasis on the range of plausible values of associations as indicated by confidence intervals [27].

## Results

### Descriptive characteristics

Characteristics of study participants (*n*=10,619; 50.3% boys) are shown in supplementary **Table S1** and weighted sample characteristics are provided in **Table S2**. The mean sample age was 14y and 35.2% of the weighted sample were overweight or obese. **Tables S3-S6** summarise the distributions of sleep characteristics. Distributions of sleep onset times and durations were similar between boys and girls; in both sexes the prevalent sleep onset time was 10-10:59pm (boys: 40.8%; girls: 40.8%) and the prevalent sleep duration was >9-10 hours (boys: 42.6%; girls: 41.6%). A higher proportion of boys than girls reported a sleep latency of 0-15 minutes (boys: 37.1%; girls: 29.7%) and never waking in the night (boys: 35.5%; girls: 24.1%). A higher proportion of girls than boys reported a sleep latency of ≥46 minutes (boys: 16.5%; girls: 19.3%), sometimes (boys: 14.3%; girls: 17.8%), often (boys: 6.7%; girls: 10.4%) and always (boys: 9.7%; girls: 14.8%) waking in the night. **Tables S7-S12** document the inter-relations between pairs of sleep characteristics.

### Associations with BMI z-score and percent body fat

There was consistent evidence for effect medication by sex hence all analyses were stratified by boys and girls (*p*≤0.0038). Results from initial models are provided in **Tables S13-S16**. Initial and model 1 results were comparable, indicating that physical activity, TV viewing, and diet did not confound or mediate associations. Patterns of association were consistent regardless of whether BMI z-score or percent body fat were modelled as outcomes. **Table 1** shows that sleep onset times after 10pm were associated with higher adiposity; associations were dose-dependent in boys but for girls plateaued after 11pm. Sleeping fewer than >9-10 hours was dose-dependently associated with higher adiposity in boys and girls, though adjustment for sleep latency and night waking attenuated associations somewhat in girls (**Table 2**). There was some evidence in both sexes that compared to a sleep latency of 16-30 minutes, a shorter latency time of 0-15 minutes was associated with higher adiposity; sleep latencies ≥46 minutes were associated with higher adiposity in girls (**Table 3**). Relative to never waking in the night and struggling to get back to sleep, sometimes waking was associated with higher adiposity in boys, and often and habitually waking were associated with higher adiposity in girls (**Table 4**). All results were substantively unchanged when instead of TV viewing, models were adjusted for social media use or electronic gaming (**Tables S17-S20**), and when percent body fat estimated by bioimpedance was modelled as the outcome (**Tables S21-S24**). Model 2 results for sleep latency and night waking frequency were consistent regardless of whether they were adjusted for sleep duration or onset time.

**Table 1.**
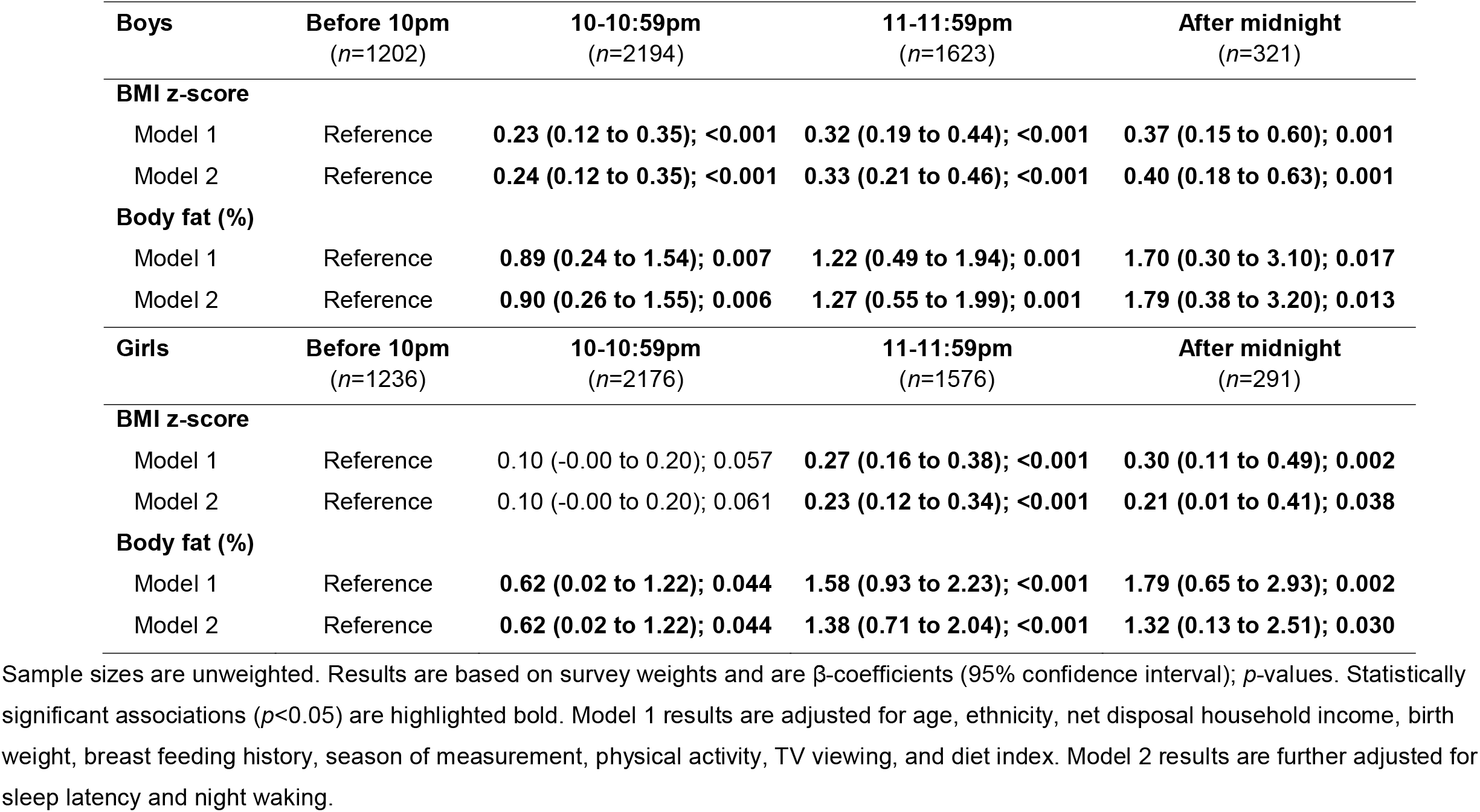
Associations of sleep onset time with adiposity.

| **Boys** | **Before 10pm**<br>(n=1202) | **10-10:59pm**<br>(n=2194) | **11-11:59pm**<br>(n=1623) | **After midnight**<br>(n=321) |
| --- | --- | --- | --- | --- |
| **BMI z-score** |  |  |  |  |
| Model 1 | Reference | **0.23 (0.12 to 0.35); <0.001** | **0.32 (0.19 to 0.44); <0.001** | **0.37 (0.15 to 0.60); 0.001** |
| Model 2 | Reference | **0.24 (0.12 to 0.35); <0.001** | **0.33 (0.21 to 0.46); <0.001** | **0.40 (0.18 to 0.63); 0.001** |
| **Body fat (%)** |  |  |  |  |
| Model 1 | Reference | **0.89 (0.24 to 1.54); 0.007** | **1.22 (0.49 to 1.94); 0.001** | **1.70 (0.30 to 3.10); 0.017** |
| Model 2 | Reference | **0.90 (0.26 to 1.55); 0.006** | **1.27 (0.55 to 1.99); 0.001** | **1.79 (0.38 to 3.20); 0.013** |
| **Girls** | **Before 10pm**<br>(n=1236) | **10-10:59pm**<br>(n=2176) | **11-11:59pm**<br>(n=1576) | **After midnight**<br>(n=291) |
| **BMI z-score** |  |  |  |  |
| Model 1 | Reference | 0.10 (-0.00 to 0.20); 0.057 | **0.27 (0.16 to 0.38); <0.001** | **0.30 (0.11 to 0.49); 0.002** |
| Model 2 | Reference | 0.10 (-0.00 to 0.20); 0.061 | **0.23 (0.12 to 0.34); <0.001** | **0.21 (0.01 to 0.41); 0.038** |
| **Body fat (%)** |  |  |  |  |
| Model 1 | Reference | **0.62 (0.02 to 1.22); 0.044** | **1.58 (0.93 to 2.23); <0.001** | **1.79 (0.65 to 2.93); 0.002** |
| Model 2 | Reference | **0.62 (0.02 to 1.22); 0.044** | **1.38 (0.71 to 2.04); <0.001** | **1.32 (0.13 to 2.51); 0.030** |

Sample sizes are unweighted. Results are based on survey weights and are $\beta$ -coefficients (95% confidence interval); $p$ -values. Statistically significant associations ( $p<0.05$ ) are highlighted bold. Model 1 results are adjusted for age, ethnicity, net disposal household income, birth weight, breast feeding history, season of measurement, physical activity, TV viewing, and diet index. Model 2 results are further adjusted for sleep latency and night waking.

**Table 2.**
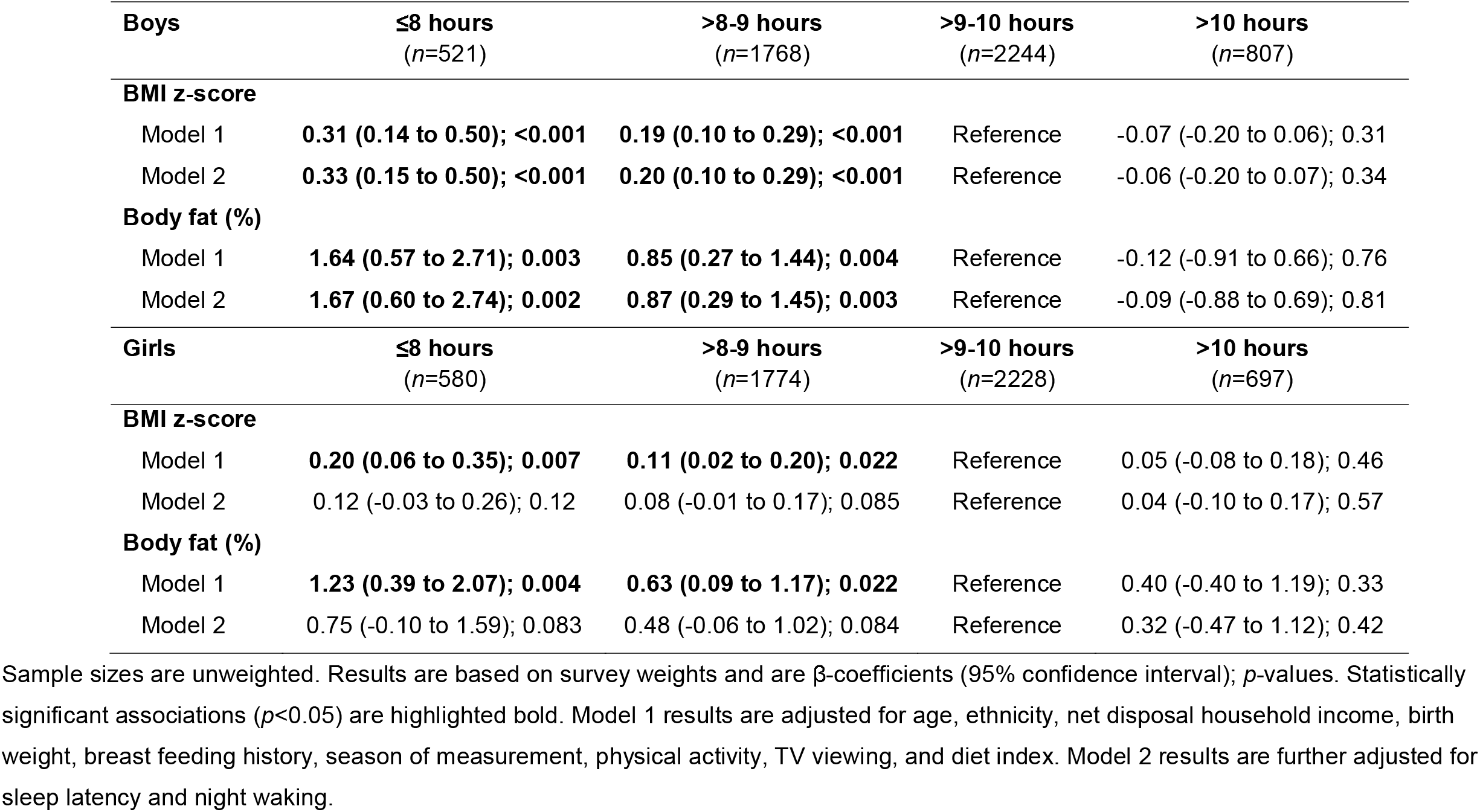
Associations of sleep duration with adiposity.

| **Boys** | **≤8 hours**<br>**(n=521)** | **>8-9 hours**<br>**(n=1768)** | **>9-10 hours**<br>**(n=2244)** | **>10 hours**<br>**(n=807)** |
| --- | --- | --- | --- | --- |
| **BMI z-score** |  |  |  |  |
| Model 1 | **0.31 (0.14 to 0.50); <0.001** | **0.19 (0.10 to 0.29); <0.001** | Reference | -0.07 (-0.20 to 0.06); 0.31 |
| Model 2 | **0.33 (0.15 to 0.50); <0.001** | **0.20 (0.10 to 0.29); <0.001** | Reference | -0.06 (-0.20 to 0.07); 0.34 |
| **Body fat (%)** |  |  |  |  |
| Model 1 | **1.64 (0.57 to 2.71); 0.003** | **0.85 (0.27 to 1.44); 0.004** | Reference | -0.12 (-0.91 to 0.66); 0.76 |
| Model 2 | **1.67 (0.60 to 2.74); 0.002** | **0.87 (0.29 to 1.45); 0.003** | Reference | -0.09 (-0.88 to 0.69); 0.81 |
| **Girls** | **≤8 hours**<br>**(n=580)** | **>8-9 hours**<br>**(n=1774)** | **>9-10 hours**<br>**(n=2228)** | **>10 hours**<br>**(n=697)** |
| **BMI z-score** |  |  |  |  |
| Model 1 | **0.20 (0.06 to 0.35); 0.007** | **0.11 (0.02 to 0.20); 0.022** | Reference | 0.05 (-0.08 to 0.18); 0.46 |
| Model 2 | 0.12 (-0.03 to 0.26); 0.12 | 0.08 (-0.01 to 0.17); 0.085 | Reference | 0.04 (-0.10 to 0.17); 0.57 |
| **Body fat (%)** |  |  |  |  |
| Model 1 | **1.23 (0.39 to 2.07); 0.004** | **0.63 (0.09 to 1.17); 0.022** | Reference | 0.40 (-0.40 to 1.19); 0.33 |
| Model 2 | 0.75 (-0.10 to 1.59); 0.083 | 0.48 (-0.06 to 1.02); 0.084 | Reference | 0.32 (-0.47 to 1.12); 0.42 |

Sample sizes are unweighted. Results are based on survey weights and are $\beta$ -coefficients (95% confidence interval); $p$ -values. Statistically significant associations ( $p < 0.05$ ) are highlighted bold. Model 1 results are adjusted for age, ethnicity, net disposal household income, birth weight, breast feeding history, season of measurement, physical activity, TV viewing, and diet index. Model 2 results are further adjusted for sleep latency and night waking.

**Table 3.** Associations of sleep latency with adiposity.

| **Boys** | **0-15 mins**<br>**(n=1989)** | **16-30 mins**<br>**(n=1743)** | **31-45 mins**<br>**(n=770)** | **46-60 mins**<br>**(n=361)** | **>60 mins**<br>**(n=477)** |
| --- | --- | --- | --- | --- | --- |
| **BMI z-score** |  |  |  |  |  |
| Model 1 | **0.11 (0.00 to 0.21); 0.042** | Reference | 0.04 (-0.09 to 0.18); 0.55 | -0.12 (-0.34 to 0.11); 0.31 | 0.04 (-0.14 to 0.23); 0.64 |
| Model 2 | **0.11 (0.01 to 0.22); 0.031** | Reference | 0.03 (-0.11 to 0.17); 0.67 | -0.15 (-0.38 to 0.08); 0.21 | -0.04 (-0.25 to 0.16); 0.66 |
| **Body fat (%)** |  |  |  |  |  |
| Model 1 | 0.54 (-0.09 to 1.16); 0.091 | Reference | 0.32 (-0.50 to 1.15); 0.44 | -0.86 (-2.21 to 0.48); 0.21 | 0.38 (-0.78 to 1.54); 0.52 |
| Model 2 | 0.59 (-0.03 to 1.21); 0.063 | Reference | 0.26 (-0.58 to 1.10); 0.54 | -1.03 (-2.40 to 0.34); 0.14 | -0.08 (-1.33 to 1.18); 0.90 |
| **Girls** | **0-15 mins**<br>**(n=1585)** | **16-30 mins**<br>**(n=1789)** | **31-45 mins**<br>**(n=909)** | **46-60 mins**<br>**(n=451)** | **>60 mins**<br>**(n=545)** |
| **BMI z-score** |  |  |  |  |  |
| Model 1 | 0.09 (-0.01 to 0.19); 0.090 | Reference | 0.09 (-0.03 to 0.22); 0.14 | **0.32 (0.17 to 0.47); <0.001** | **0.31 (0.17 to 0.45); <0.001** |
| Model 2 | 0.10 (-0.00 to 0.21); 0.061 | Reference | 0.06 (-0.06 to 0.19); 0.32 | **0.26 (0.11 to 0.41); 0.001** | **0.22 (0.08 to 0.37); 0.003** |
| **Body fat (%)** |  |  |  |  |  |
| Model 1 | 0.51 (-0.09 to 1.10); 0.094 | Reference | 0.58 (-0.13 to 1.30); 0.11 | **1.84 (0.94 to 2.74); <0.001** | **1.68 (0.85 to 2.52); <0.001** |
| Model 2 | 0.58 (-0.03 to 1.20); 0.062 | Reference | 0.39 (-0.33 to 1.11); 0.29 | **1.47 (0.57 to 2.36); 0.001** | **1.12 (0.27 to 1.97); 0.010** |

Sample sizes are unweighted. Results are based on survey weights and are $\beta$ -coefficients (95% confidence interval); $p$ -values. Statistically significant associations ( $p < 0.05$ ) are highlighted bold. Model 1 results are adjusted for age, ethnicity, net disposal household income, birth weight, breast feeding history, season of measurement, physical activity, TV viewing, and diet index. Model 2 results are further adjusted for sleep duration and night waking.

**Table 4.** Associations of night waking frequency with adiposity.

| Boys | Never<br>(n=1919) | A little<br>(n=1868) | Sometimes<br>(n=769) | Often<br>(n=345) | Habitually<br>(n=439) |
| --- | --- | --- | --- | --- | --- |
| **BMI z-score** |  |  |  |  |  |
| Model 1 | Reference | -0.01 (-0.12 to 0.10); 0.91 | 0.13 (-0.01 to 0.26); 0.078 | 0.08 (-0.10 to 0.25); 0.38 | 0.10 (-0.09 to 0.28); 0.32 |
| Model 2 | Reference | 0.00 (-0.11 to 0.11); 0.98 | **0.15 (0.01 to 0.30); 0.035** | 0.09 (-0.08 to 0.27); 0.31 | 0.10 (-0.09 to 0.30); 0.29 |
| **Body fat (%)** |  |  |  |  |  |
| Model 1 | Reference | -0.05 (-0.73 to 0.63); 0.89 | 0.75 (-0.08 to 1.58); 0.078 | 0.43 (-0.69 to 1.56); 0.45 | 0.60 (-0.50 to 1.69); 0.29 |
| Model 2 | Reference | -0.01 (-0.68 to 0.66); 0.99 | **0.88 (0.03 to 1.72); 0.043** | 0.51 (-0.62 to 1.63); 0.38 | 0.62 (-0.51 to 1.74); 0.28 |
| Girls | Never<br>(n=1313) | A little<br>(n=1814) | Sometimes<br>(n=914) | Often<br>(n=526) | Habitually<br>(n=712) |
| **BMI z-score** |  |  |  |  |  |
| Model 1 | Reference | -0.01 (-0.13 to 0.10); 0.83 | 0.08 (-0.04 to 0.21); 0.19 | **0.29 (0.14 to 0.44); <0.001** | **0.20 (0.04 to 0.36); 0.016** |
| Model 2 | Reference | -0.01 (-0.13 to 0.11); 0.87 | 0.07 (-0.06 to 0.19); 0.31 | **0.24 (0.08 to 0.41); 0.004** | 0.14 (-0.03 to 0.31); 0.12 |
| **Body fat (%)** |  |  |  |  |  |
| Model 1 | Reference | -0.13 (-0.81 to 0.54); 0.70 | 0.51 (-0.20 to 1.23); 0.16 | **1.69 (0.75 to 2.62); <0.001** | **1.30 (0.34 to 2.26); 0.008** |
| Model 2 | Reference | -0.11 (-0.81 to 0.58); 0.75 | 0.43 (-0.30 to 1.17); 0.25 | **1.44 (0.44 to 2.44); 0.005** | 0.96 (-0.05 to 1.97); 0.062 |

Sample sizes are unweighted. Results are based on survey weights and are $\beta$ -coefficients (95% confidence interval); $p$ -values. Statistically significant associations ( $p<0.05$ ) are highlighted bold. Model 1 results are adjusted for age, ethnicity, net disposal household income, birth weight, breast feeding history, season of measurement, physical activity, TV viewing, and diet index. Model 2 results are further adjusted for sleep duration and sleep latency.

### Associations with weight status

Odds ratios for overweight and obesity are shown in **Figure 1**. Compared to a sleep onset time before 10pm, going to sleep between 11-11:59pm predicted a higher likelihood of overweight and obesity in girls (1.36 (1.17 to 1.65); *p*=0.002). Associations were dose-dependent in boys for whom going to sleep after midnight was related to the highest likelihood of overweight and obesity (1.76 (1.19 to 2.60); *p*=0.004). Relative to sleeping for >9-10 hours, >8-9 hours of sleep was associated with higher odds of overweight and obesity in boys (1.38 (1.17 to 1.63); *p*<0.001) and sleeping ≤8 hours predicted higher odds of overweight and obesity in both sexes, though the association was stronger in boys than girls (boys: 1.80 (1.38 to 2.35); *p*<0.001); girls: 1.38 (1.06 to 1.79); *p*=0.016). There was evidence of a U-shaped association in girls as >10 hours of sleep was also associated with higher likelihood of overweight and obesity (1.31 (1.06 to 1.62); *p*=0.014). Compared to a sleep latency of 16-30 minutes, a sleep latency of ≥46 minutes was associated with higher likelihood of overweight and obesity in girls (46-60 minutes: 1.39 (1.05 to 1.83); *p*=0.020; >60 minutes: 1.39 (1.08 to 1.78); *p*=0.011). Full results of the logistic regression analysis are available in **Tables S25-S28**. All results were substantively unchanged when, instead of TV viewing, models were adjusted for social media use or electronic gaming (**Tables S29-S32**). Model 2 results for sleep latency and night waking frequency were consistent regardless of whether they were adjusted for sleep duration or onset time.

**Figure 1.**
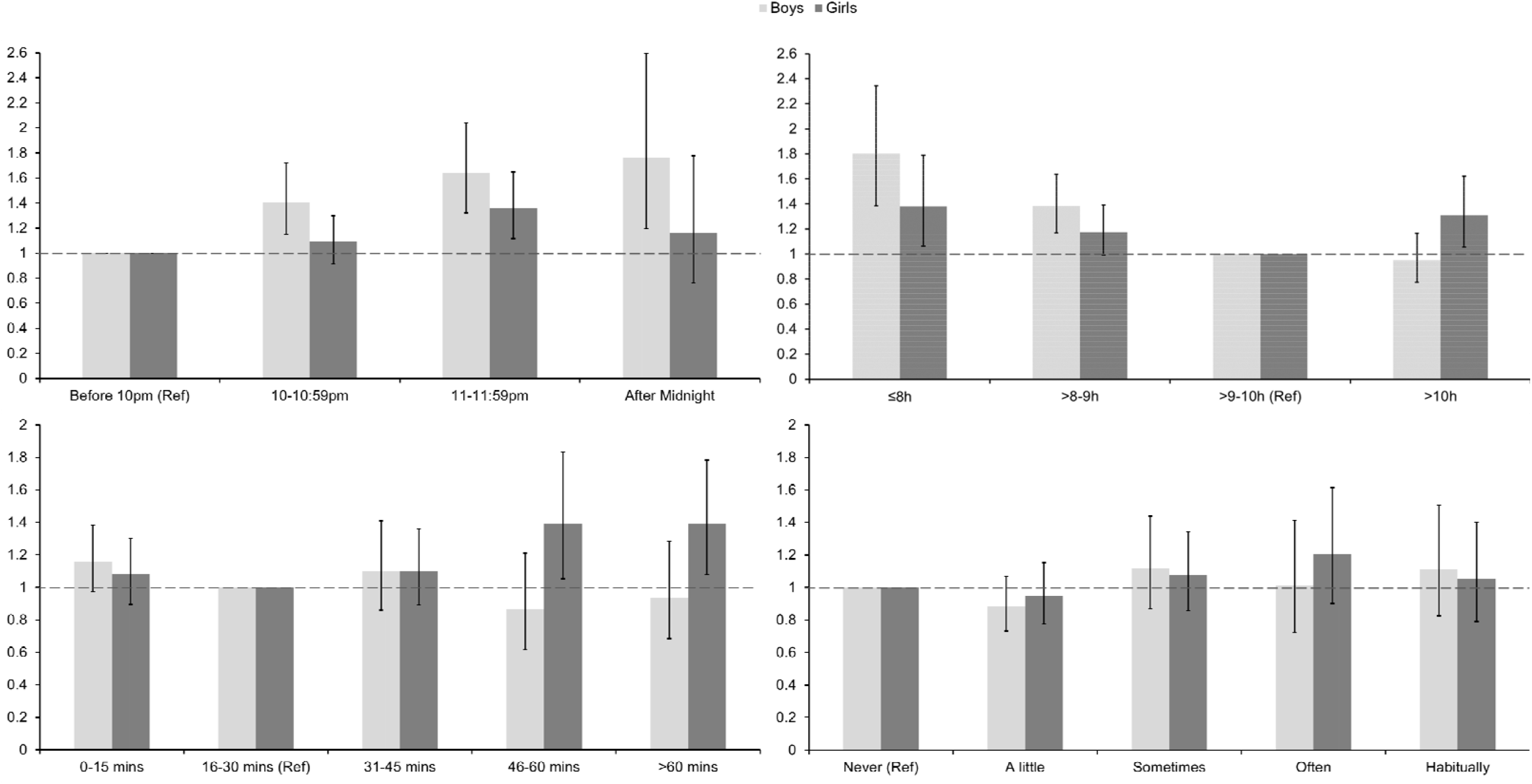
Odds ratios for overweight and obesity by sleep onset time (top left), sleep duration (top right), sleep latency (bottom left), and night waking frequency (bottom right). Results are adjusted for age, ethnicity, net disposal household income, birth weight, breast feeding history, season of measurement, physical activity, TV viewing, and diet index. Results for sleep onset time and sleep duration are further adjusted for sleep latency and night waking frequency; results for sleep latency are further adjusted for sleep duration and night waking frequency; results for night waking frequency are further adjusted for sleep duration and sleep latency.

## Discussion

This investigation of >10,000 UK adolescents has replicated the consistent finding that short sleep duration is associated with higher adiposity in youth. There was also novel evidence that sleeping longer than recommended was associated with higher odds of overweight and obesity in girls. Later sleep timing was associated with higher adiposity in both sexes, whereas sleep quality markers were consistently associated with higher adiposity and higher likelihood of overweight and obesity in girls. The results show that sleep timing, duration and quality exhibit independent associations with adolescent adiposity levels and highlight potentially important differences in relationships between boys and girls. Without exception, associations were independent of covariates including candidate mediators such as physical activity, screen time, and diet.

There is considerable evidence for associations between short sleep duration with higher expressions of BMI and obesity in youth. There is also evidence for effect modification by sex with associations appearing to be stronger in boys than girls [4, 5]. This may relate to the physiological changes that occur during puberty [28], or girls may have a lower sleep need than boys and may be more resilient to sleep debt [29]. This study also found that associations between shorter sleep duration with higher adiposity were stronger in boys than girls. Furthermore, whilst associations for boys were consistent regardless of the level of statistical adjustment, magnitudes of associations were somewhat attenuated in girls when adjusted for sleep latency and night waking frequency. In finally adjusted models, less than 8 hours of sleep was independently associated with 80% and 38% higher odds of overweight and obesity in boys and girls, respectively. These estimates are consistent with the summary odds and risk ratios produced by various meta-analyses to date, which have provided estimates ranging from 30% (1.11 to 1.53) to more than twofold (2.23; 2.18 to 2.27) higher likelihood of overweight and obesity as a function of short sleep [5]. Rather than U-shaped associations which are prevalent in adults, studies of youth report that sleep duration is linearly associated with adiposity. It is important to consider, however, that tests of non-linearity are often limited in studies of children and adolescents by low variability in sleep and sparse data at the upper end of the sleep length distribution [30]. This large national study provides novel evidence of a U-shaped association in adolescent girls, for whom longer sleep (>10 hours) was independently associated with 31% higher likelihood of overweight and obesity. This could be perceived to support the hypothesis that girls have a lower sleep need than boys, which if exceeded is detrimental. It might be that associations are also U-shaped in boys but few exceed the higher threshold above which sleep may be deleterious [31]. Depression and underlying illness are believed to contribute to the adverse health outcomes associated with long sleep in adults [32], and displacement of health-enhancing behaviours such as physical activity may also be involved. The same factors are likely implicated here. Adolescent girls experience more symptoms of anxiety and depression than boys [33], and a previous study found that adjustment for depression attenuated an association (which was initially stronger in girls) between insomnia and obesity [34]. Future studies should aim to generate more precise exposure-response curves to interrogate the shape of associations between sleep duration with adiposity and weight status in adolescents, including the potential effects of sleeping more than recommended.

A small number of studies have reported that later bed and sleep onset times are independently associated with higher BMI z-score [7, 8], waist circumference and percent body fat [8], overweight and obesity [7, 9], and obesity severity in adolescents [10]. This is the first study to report sex-specific associations in a large and nationally representative sample of adolescents. Later sleep onset was dose-dependently associated with higher adiposity in boys, for whom going to sleep after midnight predicted 76% higher odds of overweight and obesity. Magnitudes of association were somewhat attenuated in girls when adjusted for sleep latency and night waking frequency, but later sleep onset predicted higher adiposity, and going to sleep between 11-11:59pm predicted 36% higher likelihood of overweight and obesity. The best available evidence remains of low quality but suggests that short sleep, later onset, and poor sleep quality may elevate obesity risk via shared pathways, including: (a) increased neural activation in regions of the brain that are associated with food desirability, which drives a preference for nutritionally poor calorific foodstuffs, and is amplified by increased opportunity to eat due to being awake longer, (b) decreased insulin sensitivity, (c) disrupted meal timing, for instance skipping breakfast then overcompensating by eating more energy-dense foods later in the day, and (d) increased sedentary behaviour due to being awake longer (particularly more screen time late on evenings which is associated with increased snacking behaviour), and tiredness dissuading physical activity [14, 15]. The results of this study were however independent of physical activity, myriad screen-based behaviours (including TV viewing, electronic gaming, and social media use), and a composite healthy diet index. It is a weakness that all covariables were self-reported; random error and biased responses mean that residual confounding or mediation of the reported associations is likely. It is also unfortunate that only global screen time data were available, there was no time-specific information to differentiate between screen viewing on evenings compared to other times of day. Furthermore, it is a limitation that information was not available for potentially key mediating variables such as unhealthy snacking and meal times. Sleep restriction studies report increased snacking under short sleep conditions, particularly when participants would normally be asleep [35, 36], and an investigation of young children found that an association of later sleep onset with higher adiposity was attenuated when adjusted for timing of the evening meal [37]. Animal models and clinical trials of adult populations have further shown that early and time-restricted feeding has numerous physiological benefits, including lower adiposity [38, 39]. Experimental studies of sleep manipulation, and well-designed tests of mediation in observational studies which have measured variables more precisely, are needed to better shed light on the conceivable pathways of action that link sleep characteristics with obesity in youth.

Sleep quality is a heterogeneous construct and as such there is considerable variability in how it is measured and defined [11]. In this investigation, sleep latency and night waking frequency, two recognised indicators of sleep quality, were investigated. According to the National Sleep Foundation, sleep latencies of 0-15 minutes and 16-30 minutes are indicative of good sleep, whereas ≥46 minutes is a sign of poor sleep quality in youth [11]. Compared to the reference category of 16-30 minutes used in this study, sleep latencies ≥46 minutes were associated with higher adiposity and 39% higher odds of overweight and obesity in girls. This estimate is very similar to the results of a meta-analysis which found that poor sleep quality (defined broadly as longer sleep latency, more sleep disturbances, recurrent night waking, or lower sleep efficiency) predicted 46% (1.24 to 1.72) higher odds of overweight and obesity [12]. In this study there was also evidence that a short sleep latency of 0-15 minutes, which could be symptomatic of sleep deprivation, was associated with higher adiposity in both sexes. Sometimes waking in the night was associated with higher adiposity in boys and often (and to a lesser extent habitually) waking predicted higher adiposity in girls. The results suggest that poor sleep quality is associated with higher adiposity and elevated obesity risk particularly in adolescent girls, a higher proportion of whom reported longer sleep latencies and more frequent night waking than boys. Girls drink more hot caffeinated drinks in the evening [13], experience more interpersonal stress and more symptoms of anxiety and depression [33], and use their mobile phones more throughout the night than boys [41, 42]. All are potential risk factors for poor sleep quality [22].

Sleep recommendations for youth have historically focussed on duration as opposed to any other dimension of sleep [43]. The results of this study highlight that contemporary sleep guidelines should further acknowledge the importance of sleep timing and quality. Based on the results presented here, the optimal sleep pattern for 13-15y olds in terms of body composition appears to be a sleep onset time before 10pm, taking in the region of 16-45 minutes to fall asleep, and sleeping for >9-10 hours with few-to-no periods of overnight waking. Satisfying each of these conditions amidst a prevailing backdrop of short sleep may be challenging. Nonetheless, it is reassuring that associations for sleep components were independent of one another, meaning that improving just one aspect of sleep could translate to meaningful benefit. This is important given that, for example, school-based sleep education programmes are generally ineffectual in changing sleep duration but sleep quality improvements are achievable [44]. Tailoring of sleep interventions to the specific needs of boys and girls may be warranted.

### Strengths and limitations

This investigation benefitted from contemporary data collected in a nationally representative and large cohort which enabled associations to be quantified within sex strata. In addition to a weight-for-height proxy, percent body fat was investigated as a direct measure of adiposity and each of the analyses were adjusted for a broad range of covariables. This included mutual adjustment of sleep parameters to tease apart the independent associations of distinct sleep dimensions. That said, sleep onset time and duration were not mutually adjusted as they were closely related; at the fringes of cross-tabulations there were small and empty cells (see Table S7) and including both in the same statistical model produced signs of multicollinearity including reversals of direction of coefficients. As with any observational study, residual confounding by imperfectly measured covariables is possible as is confounding by unspecified factors. This study did not control for maturity status as most participants would have been in the advanced stages of puberty (>90% of boys and girls reported their voice had started getting deeper and they had experienced menarche, respectively) and there were missing data. Sleep characteristics were self-reported which is a limitation, but adolescents tend to have a set routine on school nights, and self-reports enabled sleep quality in the last four weeks to be documented. Objective sensor-based characterisation typically covers a much narrower timeframe and has low specificity for identifying periods of night waking [29, 45]. Owing to the cross-sectional nature of the analysis it is not possible to establish causality of the observed associations.

## Conclusions

Sleep timing and duration, and sleep quality in girls, appear to be independently associated with adiposity and weight status in adolescents. In addition to duration, sleep timing and quality components may be important modifiable determinants of youth overweight and obesity.

## Data Availability

The data are freely available to bona fide researchers from UK Data Service.

https://beta.ukdataservice.ac.uk/datacatalogue/series/series?id=2000031

## Acknowledgements

The author thanks the Millennium Cohort Study families for their time and cooperation, and is grateful to the Centre for Longitudinal Studies, University College London Institute of Education, for the use of the Millennium Cohort Study data and to the UK Data Archive and UK Data Service for making them available. However, they bear no responsibility for the analysis or interpretation of these data. Paul J Collings initiated this study, planned and performed the statistical analyses, interpreted the study findings, wrote the manuscript, approved the final version and is accountable for the work and any errors. The Millennium Cohort Study sixth sweep was funded by the ESRC (grant number ES/ K005987/1) and a consortium of government departments: Department for Education, Department of Health, Ministry of Justice, Home Office, Department for Transport, Department of Work and Pensions, Welsh Government, and Department for Employment and Learning (Northern Ireland). Paul J Collings is funded by a British Heart Foundation Immediate Postdoctoral Basic Science Research Fellowship (FS/17/37/32937) and is a member of The White Rose Child & Adolescent Sleep Research Network which is funded by a White Rose Collaboration Grant. The views expressed in this paper are those of the author and not necessarily those of any funder acknowledged here.

## Data availability statement

The author takes full responsibility for the integrity of the data and the accuracy of the analysis. The UK Millennium Cohort Study (MCS) obtained ethical approval from the National Health Service Research Ethics Committee system. Ethical approval has been obtained for all MCS surveys since the start of the study in 2001 and the data are freely available to bona fide researchers from UK Data Service (https://beta.ukdataservice.ac.uk/datacatalogue/series/series?id=2000031). Access to the data was granted to Paul J Collings after registration (Project id: 200285). Data analysis scripts for the current study are available from the corresponding author on reasonable request.

