## Supplemental Tables for "Independent associations of sleep timing, duration and quality with adiposity and weight status in a national sample of adolescents: the UK Millennium Cohort Study"

**Table S1. Participant characteristics based on observed data**

| | **Total (*n*=10,619)** | **Boys (*n*=5340)** | **Girls (*n*=5279)** |
|---|---|---|---|
| Age (y) | 14.3 ± 0.34 | 14.3 ± 0.34 | 14.2 ± 0.34 |
| Ethnicity | | | |
| White | 8439 (79.5) | 4239 (79.4) | 4200 (79.5) |
| South Asian | 1027 (9.7) | 501 (9.4) | 526 (10.0) |
| Other / Mixed | 1153 (10.8) | 600 (11.2) | 553 (10.5) |
| Net disposable household income (£/week) | 416 ± 178 | 420 ± 177 | 413 ± 178 |
| Birth weight (kg) | 3.4 ± 0.6 | 3.4 ± 0.6 | 3.3 ± 0.6 |
| Breast fed ≥3 months | 4253 (40.1) | 2122 (39.7) | 2131 (40.4) |
| MVPA (60 min/d) | | | |
| Not at all | 446 (4.2) | 190 (3.6) | 256 (4.9) |
| 1-2 days per week | 2523 (23.8) | 956 (17.9) | 1567 (29.7) |
| 3-4 days per week | 3606 (34.0) | 1694 (31.7) | 1912 (36.2) |
| 5-6 days per week | 2096 (19.7) | 1198 (22.4) | 898 (17.0) |
| Every day | 1948 (18.3) | 1302 (24.4) | 646 (12.2) |
| TV viewing (hours/d) | | | |
| ≤1 | 1375 (13.0) | 764 (14.3) | 611 (11.6) |
| 1-1.99 | 2133 (20.1) | 1161 (21.7) | 972 (18.4) |
| 2-2.99 | 2579 (24.3) | 1265 (23.7) | 1314 (24.9) |
| 3-4.99 | 2508 (23.6) | 1216 (22.8) | 1292 (24.5) |
| 5-6.99 | 1151 (10.8) | 538 (10.1) | 613 (11.6) |
| ≥7 | 873 (8.2) | 396 (7.4) | 477 (9.0) |
| Social media (hours/d) | | | |
| None | 826 (7.8) | 567 (10.6) | 259 (4.9) |
| <0.5 | 1309 (12.3) | 886 (16.6) | 423 (8.0) |
| 0.5-0.99 | 1532 (14.4) | 953 (17.9) | 579 (11.0) |
| 1-1.99 | 1811 (17.1) | 981 (18.4) | 830 (15.7) |
| 2-2.99 | 1639 (15.4) | 760 (14.2) | 879 (16.6) |
| 3-4.99 | 1493 (14.1) | 558 (10.4) | 935 (17.7) |
| 5-6.99 | 1013 (9.5) | 323 (6.1) | 690 (13.1) |
| ≥7 | 996 (9.4) | 312 (5.8) | 684 (13.0) |
| Electronic gaming (hours/d) | | | |
| None | 1986 (18.7) | 304 (5.7) | 1682 (31.9) |
| <0.5 | 1446 (13.6) | 312 (5.8) | 1134 (21.5) |
| 0.5-0.99 | 1246 (11.7) | 478 (8.9) | 768 (14.6) |
| 1-1.99 | 1617 (15.2) | 941 (17.6) | 676 (12.8) |
| 2-2.99 | 1449 (13.7) | 1031 (19.3) | 418 (7.9) |
| 3-4.99 | 1379 (13.0) | 1039 (19.5) | 340 (6.4) |
| 5-6.99 | 717 (6.8) | 590 (11.1) | 127 (2.4) |
| ≥7 | 779 (7.3) | 645 (12.1) | 134 (2.5) |
| Diet index | 5.5 ± 2.1 | 5.4 ± 2.0 | 5.5 ± 2.2 |
| BMI z-score | 0.57 ± 1.21 | 0.53 ± 1.24 | 0.61 ± 1.17 |
| Body fat (%) | 24.8 ± 8.1 | 21.3 ± 7.6 | 28.4 ± 6.9 |
| Weight status | | | |
| Normal | 6942 (65.4) | 3543 (66.4) | 3399 (64.4) |
| Overweight | 1588 (14.9) | 733 (13.7) | 855 (16.2) |
| Obese | 2089 (19.7) | 1064 (19.9) | 1025 (19.4) |

Unweighted sample. Values are *n* (proportions) or mean ± standard deviation. BMI, Body mass index. MVPA, Moderate to vigorous physical activity.

**Table S2. Sample characteristics based on survey weights**

| | Total | Boys | Girls |
|---|---|---|---|
| Sex (boys) | 52.3 | - | - |
| Age (y) | 14.3 (14.3 to 14.3) | 14.3 (14.3 to 14.3) | 14.3 (14.3 to 14.3) |
| Ethnicity | | | |
| White | 80.8 | 80.1 | 81.6 |
| South Asian | 6.7 | 6.6 | 6.8 |
| Other / Mixed | 12.5 | 13.3 | 11.6 |
| Net disposable household income (£/week) | 399 (388 to 409) | 401 (389 to 412) | 397 (385 to 408) |
| Birth weight (kg) | 3.3 (3.3 to 3.4) | 3.4 (3.4 to 3.4) | 3.3 (3.3 to 3.3) |
| Breast fed ≥3 months | 35.4 | 34.9 | 35.9 |
| MVPA (60 min/d) | | | |
| Not at all | 4.9 | 4.5 | 5.3 |
| 1-2 days per week | 23.8 | 18.3 | 29.9 |
| 3-4 days per week | 33.3 | 31.1 | 35.7 |
| 5-6 days per week | 19.1 | 21.4 | 16.5 |
| Every day | 18.9 | 24.7 | 12.6 |
| TV viewing (hours/d) | | | |
| ≤1 | 12.5 | 13.7 | 11.2 |
| 1-1.99 | 19.6 | 21.1 | 17.9 |
| 2-2.99 | 23.7 | 22.6 | 24.9 |
| 3-4.99 | 23.5 | 22.6 | 24.5 |
| 5-6.99 | 11.5 | 11.2 | 11.9 |
| ≥7 | 9.2 | 8.8 | 9.6 |
| Social media (hours/d) | | | |
| None | 7.7 | 10.5 | 4.7 |
| <0.5 | 12.1 | 16.2 | 7.6 |
| 0.5-0.99 | 13.7 | 16.9 | 10.1 |
| 1-1.99 | 16.7 | 18.2 | 15.0 |
| 2-2.99 | 15.2 | 14.4 | 16.2 |
| 3-4.99 | 14.1 | 10.4 | 18.3 |
| 5-6.99 | 10.0 | 6.6 | 13.5 |
| ≥7 | 10.5 | 6.8 | 14.6 |
| Electronic gaming (hours/d) | | | |
| None | 17.4 | 5.4 | 30.7 |
| <0.5 | 13.0 | 5.3 | 21.3 |
| 0.5-0.99 | 11.2 | 8.0 | 14.7 |
| 1-1.99 | 15.4 | 17.3 | 13.2 |
| 2-2.99 | 13.7 | 18.6 | 8.3 |
| 3-4.99 | 13.5 | 20.2 | 6.2 |
| 5-6.99 | 7.3 | 11.4 | 2.8 |
| ≥7 | 8.5 | 13.8 | 2.8 |
| Diet index | 5.3 (5.2 to 5.4) | 5.3 (5.2 to 5.4) | 5.3 (5.2 to 5.5) |
| BMI z-score | 0.59 (0.56 to 0.61) | 0.55 (0.51 to 0.59) | 0.63 (0.58 to 0.67) |
| Body fat (%) | 24.7 (24.4 to 24.9) | 21.3 (21.0 to 21.6) | 28.4 (28.1 to 28.7) |
| Weight status | | | |
| Normal | 64.8 | 65.8 | 63.8 |
| Overweight | 14.8 | 13.5 | 16.2 |
| Obese | 20.4 | 20.7 | 20.0 |

Values are proportions or mean (95% confidence interval) based on survey weights. BMI, Body mass index. MVPA, Moderate to vigorous physical activity.

**Table S3. Distributions of sleep onset times stratified by sex**

| | Before 10pm | 10–10:59pm | 11– 11:59pm | After midnight |
|---|---|---|---|---|
| Boys | 1202 (22.5); *23.1* | 2194 (41.1); *40.8* | 1623 (30.4); *29.8* | 321 (6.0); *6.3* |
| Girls | 1236 (23.4); *24.0* | 2176 (41.2); *40.8* | 1576 (29.9); *29.4* | 291 (5.5); *5.8* |

Values are unweighted sample sizes (proportions); *weighted proportions based on survey weights*. $Chi^2$ test on unweighted data: $p$=0.50; $Chi^2$ test on weighted data: $p$=0.67

**Table S4. Distributions of sleep duration stratified by sex**

| | ≤8 hours | 8-9 hours | 9-10 hours | >10 hours |
|---|---|---|---|---|
| Boys | 521 (9.8); *10.8* | 1768 (33.1); *31.6* | 2244 (42.0); *42.6* | 807 (15.1); *15.0* |
| Girls | 580 (11.0); *11.9* | 1774 (33.6); *33.8* | 2228 (42.2); *41.6* | 697 (13.2); *12.7* |

Values are unweighted sample sizes (proportions); *weighted proportions based on survey weights*. $Chi^2$ test on unweighted data: $p$=0.012; $Chi^2$ test on weighted data: $p$=0.02

**Table S5. Distributions of sleep latency stratified by sex**

| | 0-15 mins | 16-30 mins | 31-45 mins | 46-60 mins | >60 mins |
|---|---|---|---|---|---|
| Boys | 1989 (37.3); *37.1* | 1743 (32.6); *31.6* | 770 (14.4); *14.8* | 361 (6.8); *6.4* | 477 (8.9); *10.1* |
| Girls | 1585 (30.0); *29.7* | 1789 (33.9); *33.4* | 909 (17.2); *17.6* | 451 (8.6); *8.5* | 545 (10.3); *10.8* |

Values are unweighted sample sizes (proportions); *weighted proportions based on survey weights*. $Chi^2$ test on unweighted data: $p$<0.001; $Chi^2$ test on weighted data: $p$<0.001

**Table S6. Distributions of night waking frequency stratified by sex**

| | Never | A little | Sometimes | Often | Habitually |
|---|---|---|---|---|---|
| Boys | 1919 (35.9); *35.5* | 1868 (35.0); *33.8* | 769 (14.4); *14.3* | 345 (6.5); *6.7* | 439 (8.2); *9.7* |
| Girls | 1313 (24.9); *24.1* | 1814 (34.4); *32.9* | 914 (17.3); *17.8* | 526 (9.9); *10.4* | 712 (13.5); *14.8* |

Values are unweighted sample sizes (proportions); *weighted proportions based on survey weights*. $Chi^2$ test on unweighted data: $p$<0.001; $Chi^2$ test on weighted data: $p$<0.001

**Table S7. Cross-tabulation of sleep onset time and sleep duration**

| **Boys** | **≤8 hours** | **>8 to 9 hours** | **>9 to 10 hours** | **>10 hours** |
|---|---|---|---|---|
| Before 10pm | 0 (0.0); *0.0* | 110 (9.1); *8.8* | 560 (46.6); *47.9* | 532 (44.3); *43.3* |
| 10 – 10:59pm | 59 (2.7); *3.0* | 707 (32.2); *31.7* | 1170 (53.3); *54.1* | 258 (11.8); *11.2* |
| 11 – 11:59pm | 273 (16.8); *19.0* | 839 (51.7); *48.4* | 494 (30.4); *30.2* | 17 (1.1); *1.4* |
| After midnight | 189 (58.9); *58.3* | 112 (34.9); *34.5* | 20 (6.2); *7.2* | 0 (0.0); *0.0* |
| **Girls** | **≤8 hours** | **>8 to 9 hours** | **>9 to 10 hours** | **>10 hours** |
| Before 10pm | 0 (0.0); *0.0* | 80 (6.5); *6.7* | 644 (52.1); *54.2* | 512 (41.4); *39.1* |
| 10 – 10:59pm | 64 (2.9); *3.2* | 763 (35.1); *38.7* | 1180 (54.2); *51.0* | 169 (7.8); *7.1* |
| 11 – 11:59pm | 330 (20.9); *22.4* | 836 (53.1); *50.6* | 394 (25.0); *25.8* | 16 (1.0); *1.2* |
| After midnight | 186 (63.9); *70.2* | 95 (32.7); *26.4* | 10 (3.4); *3.4* | 0 (0.0); *0.0* |

Values are unweighted sample sizes (proportions); *weighted proportions based on survey weights*.

**Table S8. Cross-tabulation of sleep onset time and sleep latency**

| **Boys** | **0-15 mins** | **16-30 mins** | **31-45 mins** | **46-60 mins** | **>60 mins** |
|---|---|---|---|---|---|
| Before 10pm | 487 (40.5); *40.4* | 426 (35.4); *35.0* | 157 (13.1); *12.6* | 65 (5.4); *5.9* | 67 (5.6); *6.1* |
| 10 – 10:59pm | 851 (38.8); *38.4* | 752 (34.3); *33.6* | 331 (15.1); *16.2* | 155 (7.0); *5.9* | 105 (4.8); *5.9* |
| 11 – 11:59pm | 568 (35.0); *35.2* | 508 (31.3); *29.6* | 247 (15.2); *15.3* | 110 (6.8); *7.4* | 190 (11.7); *12.5* |
| After midnight | 83 (25.8); *26.0* | 57 (17.8); *15.5* | 35 (10.9); *10.5* | 31 (9.7); *7.3* | 115 (35.8); *40.7* |
| **Girls** | **0-15 mins** | **16-30 mins** | **31-45 mins** | **46-60 mins** | **>60 mins** |
| Before 10pm | 468 (37.9); *37.6* | 444 (35.9); *36.0* | 188 (15.2); *15.5* | 64 (5.2); *5.2* | 72 (5.8); *5.7* |
| 10 – 10:59pm | 683 (31.4); *31.1* | 813 (37.4); *36.4* | 391 (18.0); *18.5* | 171 (7.8); *7.8* | 118 (5.4); *6.2* |
| 11 – 11:59pm | 394 (25.0); *24.6* | 479 (30.4); *30.6* | 284 (18.0); *17.8* | 179 (11.4); *11.3* | 240 (15.2); *15.7* |
| After midnight | 40 (13.8); *12.2* | 53 (18.2); *15.9* | 46 (15.8); *19.5* | 37 (12.7); *12.9* | 115 (39.5); *39.5* |

Values are unweighted sample sizes (proportions); *weighted proportions based on survey weights*.

**Table S9. Cross-tabulation of sleep onset time and night waking frequency**

| **Boys** | **Never** | **A little** | **Sometimes** | **Often** | **Habitually** |
|---|---|---|---|---|---|
| Before 10pm | 411 (34.2); *35.3* | 450 (37.5); *36.5* | 177 (14.7); *14.6* | 64 (5.3); *4.7* | 100 (8.3); *8.9* |
| 10 – 10:59pm | 816 (37.2); *37.0* | 784 (35.7); *33.9* | 317 (14.5); *14.4* | 138 (6.3); *7.1* | 139 (6.3); *7.6* |
| 11 – 11:59pm | 579 (35.6); *34.1* | 561 (34.6); *34.0* | 230 (14.2); *14.2* | 110 (6.8); *6.2* | 143 (8.8); *11.5* |
| After midnight | 113 (35.2); *33.8* | 73 (22.7); *22.0* | 45 (14.0); *12.6* | 33 (10.3); *13.5* | 57 (17.8); *18.1* |
| **Girls** | **Never** | **A little** | **Sometimes** | **Often** | **Habitually** |
| Before 10pm | 357 (28.9); *29.2* | 437 (35.4); *33.3* | 208 (16.8); *17.2* | 97 (7.8); *7.7* | 137 (11.1); *12.6* |
| 10 – 10:59pm | 583 (26.8); *25.6* | 798 (36.7); *36.6* | 366 (16.8); *16.5* | 208 (9.5); *9.9* | 221 (10.2); *11.4* |
| 11 – 11:59pm | 328 (20.8); *19.8* | 500 (31.7); *28.8* | 294 (18.7); *20.6* | 185 (11.7); *12.9* | 269 (17.1); *17.9* |
| After midnight | 45 (15.5); *14.6* | 79 (27.1); *26.0* | 46 (15.8); *14.2* | 36 (12.4); *13.6* | 85 (29.2); *31.6* |

Values are unweighted sample sizes (proportions); *weighted proportions based on survey weights*.

**Table S10. Cross-tabulation of sleep duration and sleep latency**

| **Boys** | **0-15 mins** | **16-30 mins** | **31-45 mins** | **46-60 mins** | **>60 mins** |
|---|---|---|---|---|---|
| ≤8 hours | 160 (30.7); *28.8* | 128 (24.6); *22.8* | 72 (13.8); *14.2* | 42 (8.1); *9.5* | 119 (22.8); *24.7* |
| >8 to 9 hours | 673 (38.1); *37.8* | 592 (33.5); *32.8* | 222 (12.6); *13.1* | 114 (6.4); *5.5* | 167 (9.4); *10.8* |
| >9 to 10 hours | 875 (39.0); *40.0* | 730 (32.5); *31.1* | 354 (15.8); *16.1* | 143 (6.4); *5.9* | 142 (6.3); *6.9* |
| >10 hours | 281 (34.8); *33.5* | 293 (36.3); *36.9* | 122 (15.1); *14.8* | 62 (7.7); *7.4* | 49 (6.1); *7.4* |
| **Girls** | **0-15 mins** | **16-30 mins** | **31-45 mins** | **46-60 mins** | **>60 mins** |
| ≤8 hours | 120 (20.7); *19.9* | 134 (23.1); *21.0* | 92 (15.9); *17.8* | 83 (14.3); 15.0 | 151 (26.0); *26.3* |
| >8 to 9 hours | 491 (27.7); *26.8* | 585 (33.0); *33.6* | 324 (18.2); *18.1* | 165 (9.3); 9.4 | 209 (11.8); *12.1* |
| >9 to 10 hours | 719 (32.3); *33.0* | 843 (37.8); *36.6* | 386 (17.3); *17.9* | 148 (6.7); 6.4 | 132 (5.9); *6.1* |
| >10 hours | 255 (36.6); *36.0* | 227 (32.6); *33.8* | 107 (15.3); *15.5* | 55 (7.9); 6.8 | 53 (7.6); *7.9* |

Values are unweighted sample sizes (proportions); *weighted proportions based on survey weights*.

**Table S11. Cross-tabulation of sleep duration and night waking frequency**

| **Boys** | **Never** | **A little** | **Sometimes** | **Often** | **Habitually** |
|---|---|---|---|---|---|
| ≤8 hours | 155 (29.8); *26.6* | 150 (28.8); *28.7* | 86 (16.5); *16.1* | 46 (8.8); *9.6* | 84 (16.1); *19.0* |
| >8 to 9 hours | 624 (35.3); *35.3* | 645 (36.5); *35.4* | 227 (12.8); *12.7* | 127 (7.2); *7.5* | 145 (8.2); *9.1* |
| >9 to 10 hours | 857 (38.2); *38.4* | 794 (35.4); *33.9* | 313 (13.9); *14.1* | 130 (5.8); *5.9* | 150 (6.7); *7.7* |
| >10 hours | 283 (35.1); *34.5* | 279 (34.6); *33.8* | 143 (17.7); *17.0* | 42 (5.2); *5.0* | 60 (7.4); *9.7* |
| **Girls** | **Never** | **A little** | **Sometimes** | **Often** | **Habitually** |
| ≤8 hours | 93 (16.0); *14.9* | 153 (26.4); *24.0* | 99 (17.1); *17.6* | 76 (13.1); *12.8* | 159 (27.4); *30.7* |
| >8 to 9 hours | 410 (23.1); *22.4* | 616 (34.7); *32.7* | 327 (18.4); *18.6* | 194 (11.0); *12.8* | 227 (12.8); *13.5* |
| >9 to 10 hours | 604 (27.1); *26.2* | 813 (36.5); *36.3* | 373 (16.7); *17.4* | 192 (8.6); *8.3* | 246 (11.1); *11.8* |
| >10 hours | 206 (29.5); *30.3* | 232 (33.3); *30.7* | 115 (16.5); *16.8* | 64 (9.2); *9.4* | 80 (11.5); *12.8* |

Values are unweighted sample sizes (proportions); *weighted proportions based on survey weights*.

**Table S12. Cross-tabulation of sleep latency and night waking frequency**

| **Boys** | **Never** | **A little** | **Sometimes** | **Often** | **Habitually** |
|---|---|---|---|---|---|
| 0-15 mins | 935 (47.0); *45.9* | 683 (34.3); *33.3* | 191 (9.6); *10.7* | 61 (3.1); *3.3* | 119 (6.0); *6.8* |
| 16-30 mins | 594 (34.1); *33.7* | 670 (38.4); *38.2* | 274 (15.7); *14.4* | 112 (6.4); *7.1* | 93 (5.4); *6.6* |
| 31-45 mins | 206 (26.8); *28.6* | 274 (35.6); *34.5* | 155 (20.1); *18.4* | 70 (9.1); *8.9* | 65 (8.4); *9.6* |
| 46-60 mins | 81 (22.4); *24.2* | 120 (33.2); *33.3* | 71 (19.7); *20.1* | 40 (11.1); *9.3* | 49 (13.6); *13.1* |
| >60 mins | 103 (21.6); *20.7* | 121 (25.4); *21.3* | 78 (16.3); *17.4* | 62 (13.0); *13.1* | 113 (23.7); *27.5* |
| **Girls** | **Never** | **A little** | **Sometimes** | **Often** | **Habitually** |
| 0-15 mins | 616 (38.9); *37.5* | 579 (36.5); *34.8* | 185 (11.7); *12.8* | 88 (5.5); *6.1* | 117 (7.4); *8.8* |
| 16-30 mins | 428 (23.9); *23.3* | 725 (40.5); *39.3* | 336 (18.8); *19.6* | 139 (7.8); *8.1* | 161 (9.0); *9.7* |
| 31-45 mins | 162 (17.8); *17.9* | 290 (31.9); *30.8* | 195 (21.5); *20.4* | 133 (14.6); *14.1* | 129 (14.2); *16.8* |
| 46-60 mins | 59 (13.1); *12.1* | 117 (25.9); *24.7* | 93 (20.6); *23.9* | 83 (18.4); *17.3* | 99 (22.0); *22.0* |
| >60 mins | 48 (8.8); *9.1* | 103 (18.9); *17.8* | 105 (19.3); *16.6* | 83 (15.2); *18.7* | 206 (37.8); *37.8* |

Values are unweighted sample sizes (proportions); *weighted proportions based on survey weights*.

**Table S13. Associations of sleep onset time with adiposity – Initial model results**

| **Boys** | **Before 10pm** (*n*=1202) | **10 to 10:59pm** (*n*=2194) | **11 to 11:59pm** (*n*=1623) | **After midnight** (*n*=321) |
|---|---|---|---|---|
| BMI z-score | Reference | **0.21 (0.10 to 0.33); <0.001** | **0.29 (0.17 to 0.41); <0.001** | **0.32 (0.09 to 0.56); 0.007** |
| Body fat (%) | Reference | **0.82 (0.17 to 1.47); 0.013** | **1.14 (0.45 to 1.83); 0.001** | **1.50 (0.05 to 2.95); 0.043** |
| **Girls** | **Before 10pm** (*n*=1236) | **10 to 10:59pm** (*n*=2176) | **11 to 11:59pm** (*n*=1576) | **After midnight** (*n*=291) |
| BMI z-score | Reference | **0.11 (0.01 to 0.21); 0.032** | **0.29 (0.18 to 0.40); <0.001** | **0.33 (0.14 to 0.52); 0.001** |
| Body fat (%) | Reference | **0.68 (0.09 to 1.28); 0.024** | **1.71 (1.06 to 2.35); <0.001** | **1.96 (0.83 to 3.09); 0.001** |

Sample sizes are unweighted. Results are based on survey weights and are β-coefficients (95% confidence interval); *p*-values. Statistically significant associations ($p<0.05$) are highlighted bold. Results are adjusted for age, ethnicity, net disposal household income, birth weight, breast feeding history, season of measurement.

**Table S14. Associations of sleep duration with adiposity – Initial model results**

| **Boys** | **≤8 hours** (*n*=521) | **>8-9 hours** (*n*=1768) | **>9-10 hours** (*n*=2244) | **>10 hours** (*n*=807) |
|---|---|---|---|---|
| BMI z-score | **0.31 (0.13 to 0.49); 0.001** | **0.18 (0.08 to 0.28); <0.001** | Reference | -0.06 (-0.20 to 0.07); 0.35 |
| Body fat (%) | **1.65 (0.55 to 2.75); 0.003** | **0.80 (0.18 to 1.42); 0.012** | Reference | -0.11 (-0.91 to 0.70); 0.80 |
| **Girls** | **≤8 hours** (*n*=580) | **>8-9 hours** (*n*=1774) | **>9-10 hours** (*n*=2228) | **>10 hours** (*n*=697) |
| BMI z-score | **0.21 (0.07 to 0.36); 0.005** | **0.12 (0.03 to 0.21); 0.009** | Reference | 0.04 (-0.10 to 0.18); 0.55 |
| Body fat (%) | **1.32 (0.47 to 2.18); 0.003** | **0.72 (0.19 to 1.25); 0.008** | Reference | 0.35 (-0.46 to 1.17); 0.39 |

Sample sizes are unweighted. Results are based on survey weights and are β-coefficients (95% confidence interval); *p*-values. Statistically significant associations ($p<0.05$) are highlighted bold. Results are adjusted for age, ethnicity, net disposal household income, birth weight, breast feeding history, season of measurement.

**Table S15. Associations of sleep latency with adiposity – Initial model results**

| **Boys** | **0-15 mins** (*n*=1989) | **16-30 mins** (*n*=1743) | **31-45 mins** (*n*=770) | **46-60 mins** (*n*=361) | **>60 mins** (*n*=477) |
|---|---|---|---|---|---|
| BMI z-score | **0.10 (0.00 to 0.21); 0.047** | Reference | 0.05 (-0.09 to 0.19); 0.46 | -0.11 (-0.34 to 0.11); 0.33 | 0.04 (-0.15 to 0.23); 0.66 |
| Body fat (%) | 0.52 (-0.11 to 1.14); 0.11 | Reference | 0.39 (-0.45 to 1.23); 0.37 | -0.84 (-2.18 to 0.50); 0.22 | 0.37 (-0.82 to 1.56); 0.54 |
| **Girls** | **0-15 mins** (*n*=1585) | **16-30 mins** (*n*=1789) | **31-45 mins** (*n*=909) | **46-60 mins** (*n*=451) | **>60 mins** (*n*=545) |
| BMI z-score | 0.07 (-0.03 to 0.17); 0.14 | Reference | 0.10 (-0.02 to 0.22); 0.11 | **0.34 (0.18 to 0.49); <0.001** | **0.33 (0.20 to 0.47); <0.001** |
| Body fat (%) | 0.43 (-0.15 to 1.02); 0.15 | Reference | 0.62 (-0.09 to 1.34); 0.088 | **1.92 (1.02 to 2.83); <0.001** | **1.82 (1.00 to 2.64); <0.001** |

Sample sizes are unweighted. Results are based on survey weights and are β-coefficients (95% confidence interval); *p*-values. Statistically significant associations ($p<0.05$) are highlighted bold. Results are adjusted for age, ethnicity, net disposal household income, birth weight, breast feeding history, season of measurement.

**Table S16. Associations of night waking frequency with adiposity – Initial model results**

| **Boys** | **Never** (*n*=1919) | **A little** (*n*=1868) | **Sometimes** (*n*=769) | **Often** (*n*=345) | **Habitually** (*n*=439) |
|---|---|---|---|---|---|
| BMI z-score | Reference | 0.00 (-0.11 to 0.12); 0.95 | 0.13 (-0.01 to 0.27); 0.075 | 0.07 (-0.10 to 0.25); 0.42 | 0.07 (-0.12 to 0.27); 0.46 |
| Body fat (%) | Reference | 0.03 (-0.67 to 0.72); 0.94 | 0.77 (-0.07 to 1.61); 0.071 | 0.42 (-0.74 to 1.58); 0.48 | 0.47 (-0.66 to 1.61); 0.41 |
| **Girls** | **Never** (*n*=1313) | **A little** (*n*=1814) | **Sometimes** (*n*=914) | **Often** (*n*=526) | **Habitually** (*n*=712) |
| BMI z-score | Reference | 0.00 (-0.11 to 0.12); 0.98 | 0.10 (-0.03 to 0.22); 0.14 | **0.30 (0.15 to 0.45); <0.001** | **0.20 (0.04 to 0.36); 0.013** |
| Body fat (%) | Reference | -0.04 (-0.71 to 0.63); 0.90 | 0.60 (-0.13 to 1.32); 0.11 | **1.75 (0.83 to 2.68); <0.001** | **1.33 (0.39 to 2.26); 0.006** |

Sample sizes are unweighted. Results are based on survey weights and are β-coefficients (95% confidence interval); *p*-values. Statistically significant associations ($p<0.05$) are highlighted bold. Results are adjusted for age, ethnicity, net disposal household income, birth weight, breast feeding history, season of measurement.

**Table S17. Associations of sleep onset time with adiposity – adjusted for screen-based behaviours other than TV viewing**

| **Boys** | **Before 10pm** (*n*=1202) | **10 to 10:59pm** (*n*=2194) | **11 to 11:59pm** (*n*=1623) | **After midnight** (*n*=321) |
|---|---|---|---|---|
| **BMI z-score** | | | | |
| Model 1 + social media use | Reference | **0.23 (0.12 to 0.34); <0.001** | **0.31 (0.18 to 0.44); <0.001** | **0.36 (0.14 to 0.59); 0.002** |
| Model 2 + social media use | Reference | **0.24 (0.12 to 0.35); <0.001** | **0.33 (0.20 to 0.45); <0.001** | **0.39 (0.17 to 0.62); 0.001** |
| Model 1 + electronic games | Reference | **0.23 (0.11 to 0.35); <0.001** | **0.32 (0.19 to 0.45); <0.001** | **0.40 (0.17 to 0.62); 0.001** |
| Model 2 + electronic games | Reference | **0.23 (0.12 to 0.35); <0.001** | **0.33 (0.21 to 0.46); <0.001** | **0.43 (0.21 to 0.65); <0.001** |
| **Body fat (%)** | | | | |
| Model 1 + social media use | Reference | **0.92 (0.28 to 1.57); 0.005** | **1.28 (0.53 to 2.03); 0.001** | **1.79 (0.38 to 3.19); 0.013** |
| Model 2 + social media use | Reference | **0.94 (0.30 to 1.59); 0.004** | **1.34 (0.58 to 2.09); 0.001** | **1.89 (0.47 to 3.31); 0.009** |
| Model 1 + electronic games | Reference | **0.89 (0.22 to 1.55); 0.009** | **1.25 (0.52 to 1.98); 0.001** | **1.88 (0.50 to 3.26); 0.008** |
| Model 2 + electronic games | Reference | **0.91 (0.25 to 1.57); 0.007** | **1.32 (0.60 to 2.04); <0.001** | **1.98 (0.60 to 3.36); 0.005** |
| **Girls** | **Before 10pm** (*n*=1236) | **10 to 10:59pm** (*n*=2176) | **11 to 11:59pm** (*n*=1576) | **After midnight** (*n*=291) |
| **BMI z-score** | | | | |
| Model 1 + social media use | Reference | **0.11 (0.01 to 0.22); 0.030** | **0.28 (0.16 to 0.40); <0.001** | **0.32 (0.12 to 0.52); 0.002** |
| Model 2 + social media use | Reference | **0.11 (0.01 to 0.22); 0.031** | **0.24 (0.13 to 0.36); <0.001** | **0.23 (0.02 to 0.44); 0.029** |
| Model 1 + electronic games | Reference | **0.11 (0.01 to 0.21); 0.032** | **0.29 (0.18 to 0.40); <0.001** | **0.37 (0.18 to 0.56); <0.001** |
| Model 2 + electronic games | Reference | **0.11 (0.01 to 0.21); 0.036** | **0.25 (0.14 to 0.37); <0.001** | **0.28 (0.08 to 0.48); 0.007** |
| **Body fat (%)** | | | | |
| Model 1 + social media use | Reference | **0.72 (0.12 to 1.32); 0.019** | **1.68 (0.99 to 2.37); <0.001** | **1.96 (0.77 to 3.16); 0.001** |
| Model 2 + social media use | Reference | **0.72 (0.12 to 1.31); 0.019** | **1.48 (0.78 to 2.17); <0.001** | **1.47 (0.22 to 2.71); 0.021** |
| Model 1 + electronic games | Reference | **0.68 (0.08 to 1.28); 0.026** | **1.72 (1.07 to 2.38); <0.001** | **2.17 (1.02 to 3.32); <0.001** |
| Model 2 + electronic games | Reference | **0.67 (0.08 to 1.26); 0.027** | **1.51 (0.84 to 2.18); <0.001** | **1.68 (0.47 to 2.90); 0.007** |

Sample sizes are unweighted. Results are based on survey weights and are β-coefficients (95% confidence interval); *p*-values. Statistically significant associations ($p<0.05$) are highlighted bold. Model 1 results are adjusted for age, ethnicity, net disposal household income, birth weight, breast feeding history, season of measurement, physical activity, diet index, and social media use or electronic games as specified. Model 2 results are further adjusted for sleep latency and night waking frequency.

**Table S18. Associations of sleep duration and adiposity – adjusted for screen-based behaviours other than TV viewing**

| **Boys** | **≤8 hours** (*n*=521) | **>8 to 9 hours** (*n*=1768) | **>9 to 10 hours** (*n*=2244) | **>10 hours** (*n*=807) |
|---|---|---|---|---|
| **BMI z-score** | | | | |
| Model 1 + social media use | **0.31 (0.13 to 0.49); 0.001** | **0.20 (0.10 to 0.29); <0.001** | Reference | -0.07 (-0.20 to 0.07); 0.32 |
| Model 2 + social media use | **0.32 (0.14 to 0.50); <0.001** | **0.20 (0.11 to 0.29); <0.001** | Reference | -0.06 (-0.20 to 0.07); 0.36 |
| Model 1 + electronic games | **0.32 (0.14 to 0.50); <0.001** | **0.19 (0.10 to 0.29); <0.001** | Reference | -0.07 (-0.20 to 0.06); 0.31 |
| Model 2 + electronic games | **0.33 (0.16 to 0.51); <0.001** | **0.20 (0.10 to 0.29); <0.001** | Reference | -0.07 (-0.20 to 0.07); 0.34 |
| **Body fat (%)** | | | | |
| Model 1 + social media use | **1.68 (0.58 to 2.77); 0.003** | **0.88 (0.29 to 1.47); 0.004** | Reference | -0.14 (-0.93 to 0.64); 0.72 |
| Model 2 + social media use | **1.71 (0.62 to 2.79); 0.002** | **0.90 (0.31 to 1.48); 0.003** | Reference | -0.11 (-0.90 to 0.67); 0.78 |
| Model 1 + electronic games | **1.72 (0.62 to 2.82); 0.002** | **0.85 (0.26 to 1.45); 0.005** | Reference | -0.14 (-0.93 to 0.65); 0.73 |
| Model 2 + electronic games | **1.75 (0.66 to 2.83); 0.002** | **0.87 (0.28 to 1.46); 0.004** | Reference | -0.11 (-0.90 to 0.68); 0.78 |
| **Girls** | **≤8 hours** (*n*=580) | **>8 to 9 hours** (*n*=1774) | **>9 to 10 hours** (*n*=2228) | **>10 hours** (*n*=697) |
| **BMI z-score** | | | | |
| Model 1 + social media use | **0.19 (0.04 to 0.35); 0.012** | **0.11 (0.01 to 0.20); 0.023** | Reference | 0.04 (-0.10 to 0.18); 0.56 |
| Model 2 + social media use | 0.11 (-0.04 to 0.26); 0.16 | 0.08 (-0.01 to 0.17); 0.091 | Reference | 0.03 (-0.11 to 0.17); 0.68 |
| Model 1 + electronic games | **0.22 (0.07 to 0.37); 0.003** | **0.12 (0.03 to 0.21); 0.013** | Reference | 0.04 (-0.10 to 0.17); 0.59 |
| Model 2 + electronic games | 0.13 (-0.02 to 0.29); 0.078 | 0.09 (-0.00 to 0.18); 0.059 | Reference | 0.03 (-0.11 to 0.16); 0.70 |
| **Body fat (%)** | | | | |
| Model 1 + social media use | **1.19 (0.32 to 2.07); 0.008** | **0.63 (0.08 to 1.18); 0.025** | Reference | 0.33 (-0.48 to 1.15); 0.42 |
| Model 2 + social media use | 0.70 (-0.18 to 1.58); 0.12 | 0.47 (-0.08 to 1.02); 0.093 | Reference | 0.26 (-0.55 to 1.07); 0.53 |
| Model 1 + electronic games | **1.34 (0.47 to 2.20); 0.002** | **0.68 (0.14 to 1.22); 0.014** | Reference | 0.33 (-0.47 to 1.13); 0.42 |
| Model 2 + electronic games | 0.84 (-0.04 to 1.72); 0.060 | 0.52 (-0.02 to 1.06); 0.060 | Reference | 0.26 (-0.53 to 1.06); 0.52 |

Sample sizes are unweighted. Results are based on survey weights and are β-coefficients (95% confidence interval); *p*-values. Statistically significant associations ($p$<0.05) are highlighted bold. Model 1 results are adjusted for age, ethnicity, net disposal household income, birth weight, breast feeding history, season of measurement, physical activity, diet index, and social media use or electronic games as specified. Model 2 results are further adjusted for sleep latency and night waking frequency.

**Table S19. Associations of sleep latency and adiposity – adjusted for screen-based behaviours other than TV viewing**

| **Boys** | **0-15 mins** (*n*=1989) | **16-30 mins** (*n*=1743) | **31-45 mins** (*n*=770) | **46-60 mins** (*n*=361) | **46-60 mins** (*n*=477) |
|---|---|---|---|---|---|
| **BMI z-score** | | | | | |
| Model 1 + social media use | **0.12 (0.01 to 0.22); 0.026** | Reference | 0.04 (-0.09 to 0.18); 0.54 | -0.11 (-0.34 to 0.11); 0.33 | 0.04 (-0.14 to 0.23); 0.64 |
| Model 2 + social media use | **0.12 (0.02 to 0.22); 0.022** | Reference | 0.03 (-0.10 to 0.17); 0.64 | -0.14 (-0.37 to 0.09); 0.24 | -0.04 (-0.24 to 0.16); 0.66 |
| Model 1 + electronic games | **0.10 (0.00 to 0.21); 0.046** | Reference | 0.03 (-0.10 to 0.17); 0.62 | -0.11 (-0.32 to 0.11); 0.34 | 0.05 (-0.14 to 0.23); 0.62 |
| Model 2 + electronic games | **0.11 (0.01 to 0.22); 0.034** | Reference | 0.02 (-0.11 to 0.16); 0.74 | -0.14 (-0.36 to 0.09); 0.23 | -0.04 (-0.24 to 0.15); 0.66 |
| **Body fat (%)** | | | | | |
| Model 1 + social media use | 0.57 (-0.05 to 1.19); 0.074 | Reference | 0.33 (-0.49 to 1.15); 0.43 | -0.82 (-2.16 to 0.52); 0.23 | 0.39 (-0.78 to 1.57); 0.51 |
| Model 2 + social media use | 0.61 (-0.01 to 1.24); 0.056 | Reference | 0.26 (-0.56 to 1.09); 0.53 | -0.99 (-2.35 to 0.38); 0.16 | -0.09 (-1.35 to 1.18); 0.90 |
| Model 1 + electronic games | 0.53 (-0.10 to 1.16); 0.097 | Reference | 0.29 (-0.52 to 1.11); 0.48 | -0.81 (-2.13 to 0.51); 0.23 | 0.42 (-0.74 to 1.58); 0.48 |
| Model 2 + electronic games | 0.58 (-0.05 to 1.21); 0.069 | Reference | 0.23 (-0.60 to 1.05); 0.59 | -1.00 (-2.34 to 0.35); 0.15 | -0.06 (-1.31 to 1.19); 0.92 |
| **Girls** | **0-15 mins** (*n*=1585) | **16-30 mins** (*n*=1789) | **31-45 mins** (*n*=909) | **46-60 mins** (*n*=451) | **46-60 mins** (*n*=545) |
| **BMI z-score** | | | | | |
| Model 1 + social media use | 0.08 (-0.02 to 0.18); 0.11 | Reference | 0.10 (-0.02 to 0.22); 0.12 | **0.32 (0.17 to 0.47); <0.001** | **0.33 (0.19 to 0.47); <0.001** |
| Model 2 + social media use | 0.09 (-0.01 to 0.20); 0.077 | Reference | 0.07 (-0.05 to 0.19); 0.27 | **0.27 (0.12 to 0.42); 0.001** | **0.25 (0.10 to 0.39); 0.001** |
| Model 1 + electronic games | 0.08 (-0.02 to 0.18); 0.12 | Reference | 0.10 (-0.03 to 0.22); 0.12 | **0.32 (0.17 to 0.47); <0.001** | **0.34 (0.20 to 0.47); <0.001** |
| Model 2 + electronic games | 0.09 (-0.01 to 0.19); 0.089 | Reference | 0.07 (-0.06 to 0.19); 0.29 | **0.26 (0.11 to 0.42); 0.001** | **0.25 (0.11 to 0.39); 0.001** |
| **Body fat (%)** | | | | | |
| Model 1 + social media use | 0.47 (-0.11 to 1.05); 0.11 | Reference | 0.61 (-0.10 to 1.32); 0.094 | **1.84 (0.94 to 2.75); <0.001** | **1.77 (0.95 to 2.60); <0.001** |
| Model 2 + social media use | 0.55 (-0.05 to 1.15); 0.073 | Reference | 0.42 (-0.30 to 1.14); 0.25 | **1.49 (0.58 to 2.39); 0.001** | **1.25 (0.41 to 2.09); 0.004** |
| Model 1 + electronic games | 0.44 (-0.14 to 1.03); 0.13 | Reference | 0.58 (-0.13 to 1.30); 0.11 | **1.84 (0.94 to 2.73); <0.001** | **1.79 (0.97 to 2.60); <0.001** |
| Model 2 + electronic games | 0.51 (-0.09 to 1.12); 0.095 | Reference | 0.40 (-0.32 to 1.11); 0.28 | **1.47 (0.57 to 2.36); 0.001** | **1.23 (0.40 to 2.06); 0.004** |

Sample sizes are unweighted. Results are based on survey weights and are β-coefficients (95% confidence interval); *p*-values. Statistically significant associations ($p<0.05$) are highlighted bold. Model 1 results are adjusted for age, ethnicity, net disposal household income, birth weight, breast feeding history, season of measurement, physical activity, diet index, and social media use or electronic games as specified. Model 2 results are further adjusted for sleep duration and night waking frequency.

**Table S20. Associations of night waking frequency and adiposity – adjusted for screen-based behaviours other than TV viewing**

| **Boys** | **Never** (*n*=1919) | **A little** (*n*=1868) | **Sometimes** (*n*=769) | **Often** (*n*=345) | **Habitually** (*n*=439) |
|---|---|---|---|---|---|
| **BMI z-score** | | | | | |
| Model 1 + social media use | Reference | -0.01 (-0.12 to 0.10); 0.91 | 0.12 (-0.02 to 0.26); 0.092 | 0.07 (-0.10 to 0.24); 0.42 | 0.09 (-0.10 to 0.28); 0.36 |
| Model 2 + social media use | Reference | 0.00 (-0.11 to 0.11); 0.95 | **0.15 (0.01 to 0.29); 0.038** | 0.09 (-0.09 to 0.26); 0.33 | 0.10 (-0.09 to 0.29); 0.31 |
| Model 1 + electronic games | Reference | -0.01 (-0.12 to 0.10); 0.90 | 0.12 (-0.02 to 0.26); 0.085 | 0.08 (-0.10 to 0.25); 0.39 | 0.10 (-0.09 to 0.28); 0.31 |
| Model 2 + electronic games | Reference | 0.00 (-0.11 to 0.11); 0.99 | **0.15 (0.01 to 0.29); 0.038** | 0.09 (-0.09 to 0.27); 0.31 | 0.10 (-0.09 to 0.30); 0.29 |
| **Body fat (%)** | | | | | |
| Model 1 + social media use | Reference | -0.03 (-0.71 to 0.65); 0.94 | 0.76 (-0.07 to 1.60); 0.074 | 0.45 (-0.68 to 1.58); 0.43 | 0.60 (-0.50 to 1.69); 0.29 |
| Model 2 + social media use | Reference | 0.02 (-0.65 to 0.69); 0.95 | **0.90 (0.04 to 1.75); 0.039** | 0.52 (-0.61 to 1.66); 0.37 | 0.62 (-0.50 to 1.73); 0.28 |
| Model 1 + electronic games | Reference | -0.05 (-0.73 to 0.63); 0.88 | 0.75 (-0.09 to 1.58); 0.080 | 0.45 (-0.69 to 1.59); 0.44 | 0.61 (-0.47 to 1.70); 0.27 |
| Model 2 + electronic games | Reference | -0.01 (-0.68 to 0.66); 0.97 | **0.87 (0.02 to 1.73); 0.045** | 0.52 (-0.62 to 1.66); 0.37 | 0.62 (-0.50 to 1.74); 0.28 |
| **Girls** | **Never** (*n*=1313) | **A little** (*n*=1814) | **Sometimes** (*n*=914) | **Often** (*n*=526) | **Habitually** (*n*=712) |
| **BMI z-score** | | | | | |
| Model 1 + social media use | Reference | -0.01 (-0.12 to 0.11); 0.91 | 0.09 (-0.04 to 0.21); 0.16 | **0.30 (0.14 to 0.45); <0.001** | **0.19 (0.03 to 0.36); 0.021** |
| Model 2 + social media use | Reference | -0.01 (-0.13 to 0.11); 0.92 | 0.07 (-0.06 to 0.20); 0.30 | **0.24 (0.08 to 0.41); 0.004** | 0.12 (-0.05 to 0.29); 0.16 |
| Model 1 + electronic games | Reference | -0.01 (-0.13 to 0.10); 0.84 | 0.07 (-0.05 to 0.20); 0.25 | **0.29 (0.13 to 0.44); <0.001** | **0.20 (0.03 to 0.37); 0.019** |
| Model 2 + electronic games | Reference | -0.01 (-0.13 to 0.11); 0.84 | 0.05 (-0.08 to 0.18); 0.44 | **0.23 (0.06 to 0.40); 0.008** | 0.12 (-0.05 to 0.30); 0.16 |
| **Body fat (%)** | | | | | |
| Model 1 + social media use | Reference | -0.10 (-0.77 to 0.57); 0.77 | 0.56 (-0.16 to 1.27); 0.13 | **1.74 (0.80 to 2.67); <0.001** | **1.25 (0.28 to 2.21); 0.011** |
| Model 2 + social media use | Reference | -0.09 (-0.78 to 0.60); 0.80 | 0.45 (-0.29 to 1.20); 0.24 | **1.46 (0.46 to 2.46); 0.004** | 0.88 (-0.14 to 1.89); 0.090 |
| Model 1 + electronic games | Reference | -0.12 (-0.80 to 0.55); 0.72 | 0.46 (-0.26 to 1.19); 0.21 | **1.65 (0.69 to 2.61); 0.001** | **1.28 (0.29 to 2.26); 0.011** |
| Model 2 + electronic games | Reference | -0.12 (-0.82 to 0.58); 0.73 | 0.35 (-0.40 to 1.10); 0.36 | **1.34 (0.32 to 2.37); 0.010** | 0.88 (-0.15 to 1.92); 0.093 |

Sample sizes are unweighted. Results are based on survey weights and are β-coefficients (95% confidence interval); *p*-values. Statistically significant associations ($p<0.05$) are highlighted bold. Model 1 results are adjusted for age, ethnicity, net disposal household income, birth weight, breast feeding history, season of measurement, physical activity, diet index, and social media use or electronic games as specified. Model 2 results are further adjusted for sleep duration and sleep latency.

**Table S21. Associations of sleep onset time with percent body fat estimated by bioelectrical impedance analysis**

| **Boys** | **Before 10pm** (*n*=1187) | **10 to 10:59pm** (*n*=2168) | **11 to 11:59pm** (*n*=1595) | **After midnight** (*n*=315) |
|---|---|---|---|---|
| **Log body fat** | | | | |
| Initial model | Reference | **0.06 (0.02 to 0.10); 0.006** | **0.07 (0.02 to 0.11); 0.004** | **0.12 (0.03 to 0.20); 0.011** |
| Model 1 | Reference | **0.07 (0.02 to 0.11); 0.002** | **0.07 (0.03 to 0.12); 0.002** | **0.12 (0.04 to 0.21); 0.006** |
| Model 2 | Reference | **0.07 (0.02 to 0.11); 0.002** | **0.08 (0.03 to 0.12); 0.001** | **0.13 (0.04 to 0.21); 0.004** |
| **Girls** | **Before 10pm** (*n*=1218) | **10 to 10:59pm** (*n*=2152) | **11 to 11:59pm** (*n*=1560) | **After midnight** (*n*=289) |
| **Body fat (%)** | | | | |
| Initial model | Reference | 0.59 (0.00 to 1.19); 0.050 | **1.68 (1.02 to 2.34); <0.001** | **1.96 (0.74 to 3.18); 0.002** |
| Model 1 | Reference | 0.50 (-0.11 to 1.10); 0.11 | **1.48 (0.81 to 2.16); <0.001** | **1.71 (0.47 to 2.95); 0.007** |
| Model 2 | Reference | 0.48 (-0.12 to 1.09); 0.11 | **1.27 (0.58 to 1.95); <0.001** | 1.23 (-0.05 to 2.51); 0.059 |

Sample sizes are unweighted. Results are based on survey weights and are β-coefficients (95% confidence interval); *p*-values. Statistically significant associations ($p<0.05$) are highlighted bold. Initial model results are adjusted for age, ethnicity, net disposal household income, birth weight, breast feeding history, season of measurement. Model 1 results are further adjusted for physical activity, TV viewing, diet index. Model 2 results are further adjusted for sleep latency and night waking frequency. Due to skewness results for boys are based on log transformed percent body fat.

**Table S22. Associations of sleep duration with percent body fat estimated by bioelectrical impedance analysis**

| **Boys** | **≤8 hours** (*n*=515) | **>8 to 9 hours** (*n*=1747) | **>9 to 10 hours** (*n*=2211) | **>10 hours** (*n*=792) |
|---|---|---|---|---|
| **Log body fat** | | | | |
| Initial model | **0.07 (0.01 to 0.14); 0.028** | **0.06 (0.02 to 0.10); 0.002** | Reference | 0.01 (-0.04 to 0.06); 0.73 |
| Model 1 | **0.07 (0.00 to 0.13); 0.037** | **0.06 (0.03 to 0.10); 0.001** | Reference | 0.01 (-0.04 to 0.05); 0.82 |
| Model 2 | **0.07 (0.01 to 0.13); 0.035** | **0.06 (0.03 to 0.10); <0.001** | Reference | 0.01 (-0.04 to 0.06); 0.74 |
| **Girls** | **≤8 hours** (*n*=576) | **>8 to 9 hours** (*n*=1756) | **>9 to 10 hours** (*n*=2201) | **>10 hours** (*n*=686) |
| **Body fat (%)** | | | | |
| Initial model | **1.18 (0.28 to 2.08); 0.010** | **0.68 (0.14 to 1.22); 0.013** | Reference | 0.39 (-0.41 to 1.19); 0.34 |
| Model 1 | **1.07 (0.18 to 1.96); 0.019** | **0.59 (0.05 to 1.14); 0.033** | Reference | 0.45 (-0.33 to 1.22); 0.26 |
| Model 2 | 0.58 (-0.32 to 1.48); 0.20 | 0.42 (-0.12 to 0.97); 0.13 | Reference | 0.37 (-0.40 to 1.15); 0.34 |

Sample sizes are unweighted. Results are based on survey weights and are β-coefficients (95% confidence interval); *p*-values. Statistically significant associations ($p<0.05$) are highlighted bold. Initial model results are adjusted for age, ethnicity, net disposal household income, birth weight, breast feeding history, season of measurement. Model 1 results are further adjusted for physical activity, TV viewing, diet index. Model 2 results are further adjusted for sleep latency and night waking frequency. Due to skewness results for boys are based on log transformed percent body fat.

**Table S23. Associations of sleep latency with percent body fat estimated by bioelectrical impedance analysis**

| Boys | 0-15 mins (*n*=1964) | 16-30 mins (*n*=1709) | 31-45 mins (*n*=761) | 46-60 mins (*n*=358) | >60 mins (*n*=473) |
|---|---|---|---|---|---|
| **Log body fat** | | | | | |
| Initial model | **0.04 (0.00 to 0.07); 0.049** | Reference | 0.03 (-0.02 to 0.08); 0.21 | -0.05 (-0.13 to 0.02); 0.15 | 0.04 (-0.03 to 0.11); 0.29 |
| Model 1 | 0.04 (-0.00 to 0.07); 0.057 | Reference | 0.02 (-0.02 to 0.07); 0.33 | -0.06 (-0.14 to 0.02); 0.12 | 0.03 (-0.04 to 0.10); 0.40 |
| Model 2 | **0.04 (0.00 to 0.08); 0.039** | Reference | 0.02 (-0.03 to 0.07); 0.40 | -0.07 (-0.15 to 0.01); 0.080 | 0.01 (-0.07 to 0.08); 0.86 |
| **Girls** | **0-15 mins** (*n*=1561) | **16-30 mins** (*n*=1772) | **31-45 mins** (*n*=897) | **46-60 mins** (*n*=445) | **>60 mins** (*n*=544) |
| **Body fat (%)** | | | | | |
| Initial model | 0.32 (-0.26 to 0.90); 0.28 | Reference | 0.38 (-0.32 to 1.08); 0.29 | **1.87 (0.93 to 2.80); <0.001** | **1.86 (1.01 to 2.71); <0.001** |
| Model 1 | 0.41 (-0.18 to 1.00); 0.17 | Reference | 0.34 (-0.37 to 1.04); 0.35 | **1.74 (0.82 to 2.66); <0.001** | **1.69 (0.82 to 2.56); <0.001** |
| Model 2 | 0.50 (-0.11 to 1.11); 0.11 | Reference | 0.14 (-0.56 to 0.84); 0.69 | **1.37 (0.45 to 2.29); 0.004** | **1.17 (0.29 to 2.06); 0.010** |

Sample sizes are unweighted. Results are based on survey weights and are β-coefficients (95% confidence interval); *p*-values. Statistically significant associations ($p<0.05$) are highlighted bold. Initial model results are adjusted for age, ethnicity, net disposal household income, birth weight, breast feeding history, season of measurement. Model 1 results are further adjusted for physical activity, TV viewing, diet index. Model 2 results are further adjusted for sleep duration and night waking frequency. Due to skewness results for boys are based on log transformed percent body fat.

**Table S24. Associations of night waking frequency with percent body fat estimated by bioelectrical impedance analysis**

| **Boys** | **Never** (*n*=1893) | **A little** (*n*=1845) | **Sometimes** (*n*=756) | **Often** (*n*=338) | **Habitually** (*n*=433) |
|---|---|---|---|---|---|
| **Log body fat** | | | | | |
| Initial model | Reference | -0.02 (-0.06 to 0.03); 0.44 | 0.04 (-0.00 to 0.09); 0.068 | 0.03 (-0.03 to 0.09); 0.37 | 0.01 (-0.05 to 0.07); 0.76 |
| Model 1 | Reference | -0.02 (-0.06 to 0.03); 0.46 | **0.05 (0.00 to 0.09); 0.047** | 0.04 (-0.03 to 0.10); 0.25 | 0.02 (-0.04 to 0.08); 0.51 |
| Model 2 | Reference | -0.01 (-0.06 to 0.03); 0.54 | **0.05 (0.01 to 0.10); 0.020** | 0.04 (-0.02 to 0.10); 0.17 | 0.02 (-0.04 to 0.09); 0.46 |
| **Girls** | **Never** (*n*=1295) | **A little** (*n*=1792) | **Sometimes** (*n*=902) | **Often** (*n*=524) | **Habitually** (*n*=706) |
| **Body fat (%)** | | | | | |
| Initial model | Reference | 0.11 (-0.56 to 0.78); 0.75 | **0.76 (0.04 to 1.49); 0.040** | **2.02 (1.09 to 2.95); <0.001** | **1.29 (0.31 to 2.27); 0.010** |
| Model 1 | Reference | 0.00 (-0.68 to 0.68); 0.99 | 0.65 (-0.07 to 1.36); 0.075 | **1.94 (1.00 to 2.88); <0.001** | **1.25 (0.25 to 2.25); 0.014** |
| Model 2 | Reference | 0.02 (-0.67 to 0.71); 0.95 | 0.58 (-0.16 to 1.32); 0.13 | **1.71 (0.70 to 2.72); 0.001** | 0.93 (-0.13 to 1.99); 0.084 |

Sample sizes are unweighted. Results are based on survey weights and are β-coefficients (95% confidence interval); *p*-values. Statistically significant associations ($p<0.05$) are highlighted bold. Initial model results are adjusted for age, ethnicity, net disposal household income, birth weight, breast feeding history, season of measurement. Model 1 results are further adjusted for physical activity, TV viewing, diet index. Model 2 results are further adjusted for sleep duration and sleep latency. Due to skewness results for boys are based on log transformed percent body fat.

**Table S25. Associations of sleep onset time with weight status**

| **Boys** | **Before 10pm** (Normal weight: *n*=858 Overweight / obese: *n*=344) | **10-10:59pm** (Normal weight: *n*=1473 Overweight / obese: *n*=721) | **11-11:59pm** (Normal weight: *n*=1015 Overweight / obese: *n*=608) | **After midnight** (Normal weight: *n*=197 Overweight / obese: *n*=124) |
|---|---|---|---|---|
| **Odds ratios** | | | | |
| Initial model | Reference | **1.35 (1.11 to 1.64); 0.003** | **1.56 (1.27 to 1.91); <0.001** | **1.61 (1.11 to 2.32); 0.012** |
| Model 1 | Reference | **1.40 (1.15 to 1.72); 0.001** | **1.62 (1.30 to 2.01); <0.001** | **1.71 (1.18 to 2.48); 0.005** |
| Model 2 | Reference | **1.41 (1.15 to 1.72); 0.001** | **1.64 (1.32 to 2.04); <0.001** | **1.76 (1.19 to 2.60); 0.004** |
| **Girls** | **Before 10pm** (Normal weight: *n*=846 Overweight / obese: *n*=390) | **10-10:59pm** (Normal weight: *n*=1444 Overweight / obese: *n*=732) | **11-11:59pm** (Normal weight: *n*=935 Overweight / obese: *n*=641) | **After midnight** (Normal weight: *n*=174 Overweight / obese: *n*=117) |
| **Odds ratios** | | | | |
| Initial model | Reference | 1.12 (0.94 to 1.32); 0.21 | **1.48 (1.24 to 1.78); <0.001** | 1.36 (0.92 to 2.03); 0.13 |
| Model 1 | Reference | 1.10 (0.92 to 1.30); 0.30 | **1.43 (1.19 to 1.73); <0.001** | 1.33 (0.89 to 1.99); 0.17 |
| Model 2 | Reference | 1.09 (0.92 to 1.30); 0.33 | **1.36 (1.12 to 1.65); 0.002** | 1.16 (0.76 to 1.78); 0.49 |

Sample sizes are unweighted. Results are based on survey weights and are odds ratios (95% confidence interval); *p*-values. Statistically significant associations ($p<0.05$) are highlighted bold. Initial model results are adjusted for age, ethnicity, net disposal household income, birth weight, breast feeding history, season of measurement. Model 1 results are further adjusted for physical activity, TV viewing, diet index. Model 2 results are further adjusted for sleep latency and night waking frequency.

**Table S26. Associations of sleep duration with weight status**

| **Boys** | **≤8 hours** (Normal weight: *n*=298 Overweight / obese: *n*=223) | **>8-9 hours** (Normal weight: *n*=1128 Overweight / obese: *n*=640) | **>9-10 hours** (Normal weight: *n*=1544 Overweight / obese: *n*=700) | **>10 hours** (Normal weight: *n*=573 Overweight / obese: *n*=234) |
|---|---|---|---|---|
| **Odds ratios** | | | | |
| Initial model | **1.79 (1.39 to 2.30); <0.001** | **1.34 (1.12 to 1.59); 0.001** | Reference | 0.95 (0.77 to 1.16); 0.60 |
| Model 1 | **1.76 (1.36 to 2.29); <0.001** | **1.37 (1.15 to 1.62); <0.001** | Reference | 0.94 (0.77 to 1.15); 0.55 |
| Model 2 | **1.80 (1.38 to 2.35); <0.001** | **1.38 (1.17 to 1.63); <0.001** | Reference | 0.95 (0.77 to 1.16); 0.62 |
| **Girls** | **≤8 hours** (Normal weight: *n*=339 Overweight / obese: *n*=241) | **>8-9 hours** (Normal weight: *n*=1118 Overweight / obese: *n*=656) | **>9-10 hours** (Normal weight: *n*=1487 Overweight / obese: *n*=741) | **>10 hours** (Normal weight: *n*=455 Overweight / obese: *n*=242) |
| **Odds ratios** | | | | |
| Initial model | **1.54 (1.19 to 1.99); 0.001** | **1.24 (1.05 to 1.46); 0.012** | Reference | **1.32 (1.07 to 1.63); 0.011** |
| Model 1 | **1.52 (1.18 to 1.97); 0.001** | **1.22 (1.03 to 1.44); 0.024** | Reference | **1.33 (1.07 to 1.65); 0.009** |
| Model 2 | **1.38 (1.06 to 1.79); 0.016** | 1.17 (0.99 to 1.39); 0.065 | Reference | **1.31 (1.06 to 1.62); 0.014** |

Sample sizes are unweighted. Results are based on survey weights and are odds ratios (95% confidence interval); *p*-values. Statistically significant associations ($p<0.05$) are highlighted bold. Initial model results are adjusted for age, ethnicity, net disposal household income, birth weight, breast feeding history, season of measurement. Model 1 results are further adjusted for physical activity, TV viewing, diet index. Model 2 results are further adjusted for sleep latency and night waking frequency.

**Table S27. Associations of sleep latency with weight status**

| **Boys** | **0-15 mins** (Normal weight: *n*=1275 Overweight / obese: *n*=714) | **16-30 mins** (Normal weight: *n*=1180 Overweight / obese: *n*=563) | **31-45 mins** (Normal weight: *n*=522 Overweight / obese: *n*=248) | **46-60 mins** (Normal weight: *n*=250 Overweight / obese: *n*=111) | **>60 mins** (Normal weight: *n*=316 Overweight / obese: *n*=161) |
|---|---|---|---|---|---|
| **Odds ratios** | | | | | |
| Initial model | 1.15 (0.97 to 1.37); 0.099 | Reference | 1.14 (0.90 to 1.44); 0.29 | 0.93 (0.68 to 1.28); 0.67 | 1.11 (0.83 to 1.48); 0.48 |
| Model 1 | 1.16 (0.97 to 1.38); 0.098 | Reference | 1.11 (0.88 to 1.41); 0.37 | 0.91 (0.65 to 1.25); 0.55 | 1.08 (0.82 to 1.44); 0.58 |
| Model 2 | 1.16 (0.97 to 1.38); 0.10 | Reference | 1.10 (0.86 to 1.41); 0.44 | 0.86 (0.62 to 1.21); 0.40 | 0.94 (0.68 to 1.28); 0.69 |
| **Girls** | **0-15 mins** (Normal weight: *n*=1057 Overweight / obese: *n*=528) | **16-30 mins** (Normal weight: *n*=1178 Overweight / obese: *n*=611) | **31-45 mins** (Normal weight: *n*=583 Overweight / obese: *n*=326) | **46-60 mins** (Normal weight: *n*=267 Overweight / obese: *n*=184) | **>60 mins** (Normal weight: *n*=314 Overweight / obese: *n*=231) |
| **Odds ratios** | | | | | |
| Initial model | 1.05 (0.88 to 1.26); 0.60 | Reference | 1.14 (0.93 to 1.41); 0.20 | **1.53 (1.17 to 2.01); 0.002** | **1.57 (1.23 to 1.99); <0.001** |
| Model 1 | 1.07 (0.89 to 1.29); 0.46 | Reference | 1.14 (0.92 to 1.40); 0.24 | **1.49 (1.14 to 1.96); 0.004** | **1.54 (1.20 to 1.96); 0.001** |
| Model 2 | 1.08 (0.90 to 1.30); 0.42 | Reference | 1.10 (0.89 to 1.36); 0.37 | **1.39 (1.05 to 1.83); 0.020** | **1.39 (1.08 to 1.78); 0.011** |

Sample sizes are unweighted. Results are based on survey weights and are odds ratios (95% confidence interval); *p*-values. Statistically significant associations ($p<0.05$) are highlighted bold. Initial model results are adjusted for age, ethnicity, net disposal household income, birth weight, breast feeding history, season of measurement. Model 1 results are further adjusted for physical activity, TV viewing, diet index. Model 2 results are further adjusted for sleep duration and night waking frequency.

**Table S28. Associations of night waking frequency with weight status**

| **Boys** | **Never** (Normal weight: *n*=1267 Overweight / obese: *n*=652) | **A little** (Normal weight: *n*=1278 Overweight / obese: *n*=590) | **Sometimes** (Normal weight: *n*=495 Overweight / obese: *n*=274) | **Often** (Normal weight: *n*=233 Overweight / obese: *n*=112) | **Habitually** (Normal weight: *n*=270 Overweight / obese: *n*=169) |
|---|---|---|---|---|---|
| **Odds ratios** | | | | | |
| Initial model | Reference | 0.90 (0.75 to 1.08); 0.26 | 1.09 (0.86 to 1.39); 0.47 | 1.00 (0.72 to 1.38); 0.99 | 1.07 (0.80 to 1.43); 0.63 |
| Model 1 | Reference | 0.88 (0.73 to 1.06); 0.19 | 1.09 (0.85 to 1.40); 0.49 | 1.01 (0.73 to 1.40); 0.96 | 1.12 (0.84 to 1.49); 0.43 |
| Model 2 | Reference | 0.88 (0.73 to 1.07); 0.20 | 1.12 (0.87 to 1.44); 0.38 | 1.01 (0.72 to 1.41); 0.95 | 1.11 (0.82 to 1.50); 0.48 |
| **Girls** | **Never** (Normal weight: *n*=871 Overweight / obese: *n*=442) | **A little** (Normal weight: *n*=1218 Overweight / obese: *n*=596) | **Sometimes** (Normal weight: *n*=577 Overweight / obese: *n*=337) | **Often** (Normal weight: *n*=309 Overweight / obese: *n*=217) | **Habitually** (Normal weight: *n*=424 Overweight / obese: *n*=288) |
| **Odds ratios** | | | | | |
| Initial model | Reference | 0.98 (0.81 to 1.18); 0.81 | 1.14 (0.92 to 1.43); 0.23 | **1.35 (1.03 to 1.76); 0.030** | 1.20 (0.92 to 1.57); 0.18 |
| Model 1 | Reference | 0.95 (0.78 to 1.15); 0.60 | 1.12 (0.90 to 1.39); 0.32 | **1.32 (1.01 to 1.74); 0.046** | 1.20 (0.91 to 1.58); 0.20 |
| Model 2 | Reference | 0.95 (0.78 to 1.15); 0.59 | 1.07 (0.86 to 1.34); 0.54 | 1.21 (0.90 to 1.62); 0.21 | 1.05 (0.79 to 1.40); 0.73 |

Sample sizes are unweighted. Results are based on survey weights and are odds ratios (95% confidence interval); *p*-values. Statistically significant associations ($p<0.05$) are highlighted bold. Initial model results are adjusted for age, ethnicity, net disposal household income, birth weight, breast feeding history, season of measurement. Model 1 results are further adjusted for physical activity, TV viewing, diet index. Model 2 results are further adjusted for sleep duration and sleep latency.

**Table S29. Associations of sleep onset time with weight status – adjusted for screen-based behaviours other than TV viewing**

| **Boys** | **Before 10pm** (Normal weight: *n*=858 Overweight / obese: *n*=344) | **10-10:59pm** (Normal weight: *n*=1473 Overweight / obese: *n*=721) | **11-11:59pm** (Normal weight: *n*=1015 Overweight / obese: *n*=608) | **After midnight** (Normal weight: *n*=197 Overweight / obese: *n*=124) |
|---|---|---|---|---|
| **Odds ratios** | | | | |
| Model 1 + social media use | Reference | **1.42 (1.16 to 1.74); 0.001** | **1.66 (1.32 to 2.07); <0.001** | **1.77 (1.23 to 2.56); 0.002** |
| Model 2 + social media use | Reference | **1.43 (1.16 to 1.75); 0.001** | **1.68 (1.34 to 2.10); <0.001** | **1.82 (1.23 to 2.69); 0.003** |
| Model 1 + electronic games | Reference | **1.41 (1.15 to 1.72); 0.001** | **1.65 (1.33 to 2.04); <0.001** | **1.80 (1.26 to 2.59); 0.001** |
| Model 2 + electronic games | Reference | **1.41 (1.16 to 1.72); 0.001** | **1.67 (1.35 to 2.07); <0.001** | **1.85 (1.27 to 2.70); 0.001** |
| **Girls** | **Before 10pm** (Normal weight: *n*=846 Overweight / obese: *n*=390) | **10-10:59pm** (Normal weight: *n*=1444 Overweight / obese: *n*=732) | **11-11:59pm** (Normal weight: *n*=935 Overweight / obese: *n*=641) | **After midnight** (Normal weight: *n*=174 Overweight / obese: *n*=117) |
| **Odds ratios** | | | | |
| Model 1 + social media use | Reference | 1.15 (0.96 to 1.37); 0.12 | **1.53 (1.25 to 1.87); <0.001** | 1.46 (0.97 to 2.21); 0.073 |
| Model 2 + social media use | Reference | 1.14 (0.96 to 1.36); 0.14 | **1.45 (1.18 to 1.78); <0.001** | 1.26 (0.82 to 1.95); 0.29 |
| Model 1 + electronic games | Reference | 1.12 (0.94 to 1.33); 0.22 | **1.49 (1.24 to 1.80); <0.001** | 1.48 (0.97 to 2.25); 0.066 |
| Model 2 + electronic games | Reference | 1.11 (0.93 to 1.32); 0.25 | **1.41 (1.16 to 1.71); 0.001** | 1.28 (0.83 to 1.99); 0.27 |

Sample sizes are unweighted. Results are based on survey weights and are odds ratios (95% confidence interval); *p*-values. Statistically significant associations ($p<0.05$) are highlighted bold. Model 1 results are adjusted for age, ethnicity, net disposal household income, birth weight, breast feeding history, season of measurement, physical activity, diet index, and social media use or electronic games as specified. Model 2 results are further adjusted for sleep latency and night waking frequency.

**Table S30. Associations of sleep duration with weight status – adjusted for screen-based behaviours other than TV viewing**

| **Boys** | **≤8 hours** (Normal weight: *n*=298 Overweight / obese: *n*=223) | **>8-9 hours** (Normal weight: *n*=1128 Overweight / obese: *n*=640) | **>9-10 hours** (Normal weight: *n*=1544 Overweight / obese: *n*=700) | **>10 hours** (Normal weight: *n*=573 Overweight / obese: *n*=234) |
|---|---|---|---|---|
| **Odds ratios** | | | | |
| Model 1 + social media use | **1.81 (1.40 to 2.35); <0.001** | **1.38 (1.16 to 1.64); <0.001** | Reference | 0.94 (0.76 to 1.15); 0.54 |
| Model 2 + social media use | **1.85 (1.42 to 2.40); <0.001** | **1.39 (1.17 to 1.65); <0.001** | Reference | 0.95 (0.77 to 1.16); 0.61 |
| Model 1 + electronic games | **1.82 (1.41 to 2.36); <0.001** | **1.37 (1.15 to 1.62); <0.001** | Reference | 0.94 (0.76 to 1.15); 0.52 |
| Model 2 + electronic games | **1.86 (1.43 to 2.41); <0.001** | **1.38 (1.17 to 1.64); <0.001** | Reference | 0.95 (0.77 to 1.16); 0.59 |
| **Girls** | **≤8 hours** (Normal weight: *n*=339 Overweight / obese: *n*=241) | **>8-9 hours** (Normal weight: *n*=1118 Overweight / obese: *n*=656) | **>9-10 hours** (Normal weight: *n*=1487 Overweight / obese: *n*=741) | **>10 hours** (Normal weight: *n*=455 Overweight / obese: *n*=242) |
| **Odds ratios** | | | | |
| Model 1 + social media use | **1.55 (1.20 to 2.1); 0.001** | **1.23 (1.04 to 1.45); 0.017** | Reference | **1.28 (1.03 to 1.59); 0.024** |
| Model 2 + social media use | **1.40 (1.07 to 1.82); 0.013** | 1.18 (1.00 to 1.40); 0.050 | Reference | **1.26 (1.02 to 1.57); 0.035** |
| Model 1 + electronic games | **1.57 (1.22 to 2.03); 0.001** | **1.23 (1.04 to 1.45); 0.018** | Reference | **1.30 (1.05 to 1.61); 0.016** |
| Model 2 + electronic games | **1.41 (1.08 to 1.84); 0.010** | 1.18 (1.00 to 1.40); 0.052 | Reference | **1.28 (1.03 to 1.59); 0.023** |

Sample sizes are unweighted. Results are based on survey weights and are odds ratios (95% confidence interval); *p*-values. Statistically significant associations ($p<0.05$) are highlighted bold. Model 1 results are adjusted for age, ethnicity, net disposal household income, birth weight, breast feeding history, season of measurement, physical activity, diet index, and social media use or electronic games as specified. Model 2 results are further adjusted for sleep latency and night waking frequency.

**Table S31. Associations of sleep latency with weight status – adjusted for screen-based behaviours other than TV viewing**

| **Boys** | **0-15 mins** (Normal weight: *n*=1275 Overweight / obese: *n*=714) | **16-30 mins** (Normal weight: *n*=1180 Overweight / obese: *n*=563) | **31-45 mins** (Normal weight: *n*=522 Overweight / obese: *n*=248) | **46-60 mins** (Normal weight: *n*=250 Overweight / obese: *n*=111) | **>60 mins** (Normal weight: *n*=316 Overweight / obese: *n*=161) |
|---|---|---|---|---|---|
| **Odds ratios** | | | | | |
| Model 1 + social media use | 1.18 (0.99 to 1.40); 0.068 | Reference | 1.11 (0.88 to 1.41); 0.38 | 0.93 (0.67 to 1.28); 0.65 | 1.11 (0.84 to 1.48); 0.46 |
| Model 2 + social media use | 1.17 (0.98 to 1.40); 0.079 | Reference | 1.10 (0.86 to 1.40); 0.45 | 0.89 (0.63 to 1.24); 0.48 | 0.96 (0.70 to 1.31); 0.78 |
| Model 1 + electronic games | 1.16 (0.98 to 1.38); 0.092 | Reference | 1.11 (0.87 to 1.40); 0.40 | 0.93 (0.68 to 1.28); 0.67 | 1.11 (0.84 to 1.48); 0.47 |
| Model 2 + electronic games | 1.16 (0.97 to 1.38); 0.096 | Reference | 1.09 (0.86 to 1.39); 0.47 | 0.89 (0.64 to 1.23); 0.48 | 0.95 (0.70 to 1.31); 0.77 |
| **Girls** | **0-15 mins** (Normal weight: *n*=1057 Overweight / obese: *n*=528) | **16-30 mins** (Normal weight: *n*=1178 Overweight / obese: *n*=611) | **31-45 mins** (Normal weight: *n*=583 Overweight / obese: *n*=326) | **46-60 mins** (Normal weight: *n*=267 Overweight / obese: *n*=184) | **>60 mins** (Normal weight: *n*=314 Overweight / obese: *n*=231) |
| **Odds ratios** | | | | | |
| Model 1 + social media use | 1.06 (0.88 to 1.26); 0.55 | Reference | 1.15 (0.93 to 1.41); 0.21 | **1.51 (1.15 to 1.98); 0.003** | **1.58 (1.24 to 2.01); <0.001** |
| Model 2 + social media use | 1.06 (0.89 to 1.28); 0.51 | Reference | 1.11 (0.90 to 1.37); 0.33 | **1.40 (1.06 to 1.86); 0.017** | **1.43 (1.11 to 1.84); 0.005** |
| Model 1 + electronic games | 1.06 (0.88 to 1.27); 0.54 | Reference | 1.14 (0.93 to 1.42); 0.21 | **1.50 (1.15 to 1.97); 0.003** | **1.59 (1.24 to 2.03); <0.001** |
| Model 2 + electronic games | 1.06 (0.88 to 1.27); 0.52 | Reference | 1.11 (0.90 to 1.37); 0.33 | **1.40 (1.06 to 1.84); 0.017** | **1.43 (1.12 to 1.84); 0.005** |

Sample sizes are unweighted. Results are based on survey weights and are odds ratios (95% confidence interval); *p*-values. Statistically significant associations ($p<0.05$) are highlighted bold. Model 1 results are adjusted for age, ethnicity, net disposal household income, birth weight, breast feeding history, season of measurement, physical activity, diet index, and social media use or electronic games as specified. Model 2 results are further adjusted for sleep duration and night waking frequency.

**Table S32. Associations of night waking frequency with weight status – adjusted for screen-based behaviours other than TV viewing**

| **Boys** | **Never** (Normal weight: *n*=1267 Overweight / obese: *n*=652) | **A little** (Normal weight: *n*=1278 Overweight / obese: *n*=590) | **Sometimes** (Normal weight: *n*=495 Overweight / obese: *n*=274) | **Often** (Normal weight: *n*=233 Overweight / obese: *n*=112) | **Habitually** (Normal weight: *n*=270 Overweight / obese: *n*=169) |
|---|---|---|---|---|---|
| **Odds ratios** | | | | | |
| Model 1 + social media use | Reference | 0.88 (0.73 to 1.06); 0.19 | 1.09 (0.85 to 1.40); 0.50 | 1.00 (0.72 to 1.39); 0.99 | 1.13 (0.85 to 1.51); 0.40 |
| Model 2 + social media use | Reference | 0.89 (0.73 to 1.07); 0.21 | 1.12 (0.87 to 1.45); 0.38 | 1.00 (0.72 to 1.40); 0.99 | 1.12 (0.83 to 1.51); 0.47 |
| Model 1 + electronic games | Reference | 0.88 (0.73 to 1.06); 0.19 | 1.09 (0.84 to 1.41); 0.51 | 1.01 (0.73 to 1.41); 0.93 | 1.13 (0.85 to 1.50); 0.39 |
| Model 2 + electronic games | Reference | 0.88 (0.73 to 1.07); 0.20 | 1.11 (0.86 to 1.44); 0.41 | 1.01 (0.72 to 1.41); 0.94 | 1.12 (0.83 to 1.50); 0.47 |
| **Girls** | **Never** (Normal weight: *n*=871 Overweight / obese: *n*=442) | **A little** (Normal weight: *n*=1218 Overweight / obese: *n*=596) | **Sometimes** (Normal weight: *n*=577 Overweight / obese: *n*=337) | **Often** (Normal weight: *n*=309 Overweight / obese: *n*=217) | **Habitually** (Normal weight: *n*=424 Overweight / obese: *n*=288) |
| **Odds ratios** | | | | | |
| Model 1 + social media use | Reference | 0.96 (0.79 to 1.17); 0.71 | 1.13 (0.91 to 1.41); 0.27 | **1.36 (1.04 to 1.80); 0.026** | 1.19 (0.90 to 1.58); 0.22 |
| Model 2 + social media use | Reference | 0.96 (0.79 to 1.17); 0.67 | 1.08 (0.86 to 1.35); 0.52 | 1.23 (0.92 to 1.65); 0.16 | 1.03 (0.77 to 1.38); 0.83 |
| Model 1 + electronic games | Reference | 0.95 (0.78 to 1.15); 0.60 | 1.10 (0.88 to 1.37); 0.41 | 1.31 (1.00 to 1.73); 0.054 | 1.20 (0.90 to 1.59); 0.21 |
| Model 2 + electronic games | Reference | 0.94 (0.77 to 1.15); 0.56 | 1.04 (0.83 to 1.31); 0.71 | 1.18 (0.88 to 1.58); 0.27 | 1.04 (0.78 to 1.39); 0.80 |

Sample sizes are unweighted. Results are based on survey weights and are odds ratios (95% confidence interval); *p*-values. Statistically significant associations ($p<0.05$) are highlighted bold. Model 1 results are adjusted for age, ethnicity, net disposal household income, birth weight, breast feeding history, season of measurement, physical activity, diet index, and social media use or electronic games as specified. Model 2 results are further adjusted for sleep duration and sleep latency.
